# A conserved regulatory atlas reveals enhancers critical for mouse and human gonadal development

**DOI:** 10.64898/2026.07.09.26357422

**Authors:** Aviva Eliyahu, Ido Blass, Meshi Ridnik, Isabelle Stevant, Gabby Atlas, Roni Weiss, Elisheva Abberbock, Jocelyn van den Bergen, Gorjana Robevska, Katrina M Bell, Lucas G A Ferreira, Elena J Tucker, Brendan J Houston, Kim C Donaghue, Joel A Vanderniet, Catherine S Choong, Magnus Dias da Silva, Laurene Schlick-Garcia, Estelle Talouarn, Maeva Elzaiat, Ofer Feinstein, Michal Linial, Yaron Orenstein, Anu Bashamboo, Andrew H Sinclair, Kenneth McElreavey, Katie L Ayers, Nitzan Gonen

**Author notes:** These authors contributed equally (Aviva Eliyahu, Ido Blass, Meshi Ridnik).

## Abstract

Differences of Sex Development (DSD) are congenital conditions where sex development is atypical. Like many other rare conditions, genetic diagnostic rates for DSD still sit below 50%. Unexplained cases may harbour disruptions in non-coding regulatory regions, which remain largely unexplored. Here, we integrate chromatin accessibility datasets from mouse and human embryonic gonads to construct a complete comparative regulatory atlas of gonad development. Using this resource, we identified a conserved upstream enhancer of the key gene, *Wt1*. We identify rare variants in this enhancer in eight individuals with DSD and demonstrate that its deletion in mice results in XY male-to-female sex reversal and XX sub-fertility, thereby establishing it as critical for gonadal development. We build on this atlas to create a tissue-specific computational filtration pipeline to prioritise non-coding variants in whole-genome sequencing datasets from 74 undiagnosed individuals with DSD. This dramatically reduces millions of potential candidates to a small set of candidate variants. This led to identification of a variant that disrupts the activity a conserved enhancer upstream of *NR5A1* which segregates with gonadal dysgenesis in a family. Our study reveals regulatory elements essential for gonadal development and provides a framework for uncovering pathogenic non-coding variants in other rare developmental conditions.

## Introduction

Despite rapid improvements in genomic technologies over the past decade, the genetic diagnostic yield for many rare congenital conditions remains ∼45% ^1^. Indeed, current genetic diagnostic approaches are often limited to protein-coding regions of genes, which represent just 2% of the genome. The non-coding genome, which includes *cis*-regulatory elements (*c*REs) such as promoters, enhancers, silencers, and insulators, all of which control gene expression, are by in large not analysed. However, there is now a growing recognition that common disease-associated variants are disproportionately enriched within *c*REs ^2^ and disruptions to *c*REs have been linked to various disorders ^3^. Indeed, genome wide association studies (GWAS) indicate that a majority of disease-linked genetic variants reside in the non-coding gnome, mainly within *c*REs ^4–6^. The increased availability, and reduced cost, of whole genome sequencing (WGS), including long read approaches, now enables the identification of variants in non-coding regions in undiagnosed patients ^4,7^. However, the vast number of variants identified by these approaches presents a major challenge for validation or variant curation. Indeed, an individual will carry, on average, 4-5 million rare single nucleotide variants (SNV) and 2,100 - 2,500 structural variants compared to the reference genome ^8^. This challenge is compounded by the lack of understanding about the function of the non-coding genome, especially in some understudied human tissues, and the limited *in silico* or functional tools with which to assess non-coding variant pathogenicity. These variants are often classified as variants of uncertain significance (VUS). Despite recent efforts to provide guidance on the interpretation and functional analysis of non-coding variants ^9,10^, major challenges still exist, particularly for rare developmental conditions affecting tissues that arise early in embryogenesis, where access to relevant tissue or appropriate functional models is limited. This includes conditions where reproductive development is atypical, such as differences of sex development (DSD).

In mammals, sex is determined by inheritance of the sex chromosomes which drives the first step in reproductive development - the differentiation of an initially bipotential gonad towards a testicular or ovarian fate. This process relies on a delicate and highly regulated balance between two antagonistic genetic programs involving many genes, most of which are transcription factors and signalling pathway components ^11–13^ whose expression is tightly controlled. The bipotential gonad develops at embryonic day (E) 10.5 in mice and at around 5 gestational weeks (GW) in humans. In individuals with an XY karyotype, expression of the Y-linked gene, *SRY*, along with its direct target gene, *SOX9* within supporting cell precursors, drives their differentiation into Sertoli cells ^14–16^. Sertoli cells subsequently direct the differentiation of other gonadal somatic cell populations towards interstitial testicular cells such as hormone producing Leydig cells and germ cells to develop into spermatozoa. Studies primarily conducted in animal models have shown that ovarian differentiation relies on the absence of *Sry* and the joint activity of several partially redundant factors including *Wt1* (-KTS isoform), *Wnt4/Rspo1*, *Foxl2*, and *Runx1* (reviewed in ^17^). Several of these genes have also been shown to be involved in human sex development and therefore contribute to DSD when their function is disrupted ^11^. Expression of these factors within supporting cell precursors directs their differentiation into granulosa cells, which subsequently direct ovary and oocyte development. Once the testes and ovaries have formed, sex differentiation of the internal ductal system and external genitalia occurs, under the influence of sex hormones produced by the gonads ^18^. The mouse is considered the closest model system for human gonad development and while similarities exist, important differences are also evident ^19^.

DSD are defined as congenital conditions in which the development of chromosomal, gonadal or anatomical sex is atypical ^7,11,20,21^. DSD is an umbrella term that includes highly variable clinical phenotypes ranging from 46,XY gonadal dysgenesis with a females phenotype, to formation of ovotestis or ambiguous genitalia, to milder phenotypes such as cryptorchidism or micropenis. DSD affect 1-2% of live births ^22^ with genital anomalies accounting for 7.5% of all birth defects. Clinical care requires substantial healthcare resources often involving surgery, hormone therapy and fertility preservation, whilst maximising psychosocial well-being ^20^. An increased risk of gonadal cancer in some 46,XY DSD requires monitoring often via invasive biopsy or even prophylactic gonadectomy. Establishing a clear genetic diagnosis can substantially improve clinical care by providing diagnostic certainty, determining possible co-morbidities, informing on cancer or infertility risk, and enabling personalised management strategies ^23,24^. More than three decades of research in animal models and with individuals with DSD have highlighted many of the genes involved in gonad development, with ∼100 genes now considered diagnostic for DSD or associated conditions (<u>PanelApp</u>) ^25^. Yet, although many patients can now access exome sequencing as part of their clinical care, genetic diagnostic rates have remained at ∼45%, for 46,XY DSD and ∼30% of 46,XX DSD ^7,26,27^, strongly suggesting that additional genetic causes remain to be found, including variants located within the non-coding genome. Indeed, we have previously identified DSD patients with disruptions to enhancers responsible for testicular expression of *SRY* ^28^ or *SOX9* ^29^. We have demonstrated that some of these, notably the human *SOX9 e*SRA enhancer, known as Enh13 in mice, are functionally conserved ^30–35^. These DSD “enhanceropathies” have now been integrated into diagnostic frameworks, and for some patient groups, have delivered a 10% increase in genetic diagnoses, similar to what is seen in other conditions ^36^.

In recent years, several studies have started to map *c*REs instructive of gonadal development in both mouse and humans ^37–41^. Yet the extent of conservation between the mouse and human regulatory landscapes has not been explored. Furthermore, these novel datasets have not been exploited for the identification and functional analysis of candidate variants in undiagnosed cases of DSD.

Here we present an in-depth comparison of the mouse and human regulatory landscapes during early embryonic gonad development. We found that 36% of the regulatory elements accessible in the supporting cells of human embryonic ovaries and testes are also accessible in mouse counterparts. Focusing on *c*REs associated with key gonad developmental genes, we identified a novel upstream enhancer of the mouse *Wt1* gene and show that deletion of this enhancer leads to XY sex reversal. These XY mice are born as phenotypic females with dysgenic ovaries and reproductive systems containing both Müllerian and Wolffian derivatives, similar to previously reported individuals with DSD carrying pathogenic *WT1* variants ^42^. Analysis of this enhancer region in undiagnosed individuals with DSD identified rare heterozygous variants in eight individuals with DSD. To expand this analysis, we developed a computational filtration pipeline based on our conserved gonadal regulatory atlas to prioritise rare variants from WGS of a large cohort of undiagnosed individuals with DSD. This approach identified several candidate variants in *c*REs linked to known or suspected DSD genes. This included a variant in a conserved enhancer for *NR5A1*. The *NR5A1* gene is a key regulator of gonad development in which pathogenic variants are an established cause of DSD. This *NR5A1* enhancer variant segregated with 46,XY gonadal dysgenesis in a family and functional testing demonstrated this variant causes a reduction of enhancer activity *in vitro*, which we postulate to be through the strengthening of a binding site for a transcriptional repressor TCF3, a factor highly expressed in the testes. In summary, this study provides the first conserved regulatory atlas in developing foetal ovaries and testis, revealing novel enhancers required for gonadal differentiation and identifying candidate patient variants. Furthermore, it paves the way for solving unexplained cases of DSD while providing a framework applicable to the study of other rare congenital conditions.

## Results

### A conserved regulatory map of gonadal development

In the developing embryonic gonads, differentiation into ovaries or testes is mediated by the supporting cell precursors, which give rise to the somatic Sertoli cells in the testis or granulosa cells in the ovary. Sertoli and granulosa cells then drive differentiation of additional cell types including the steroidogenic and germ lineages. The differentiation into Sertoli or granulosa cells takes place between E11.5 and E13.5 in mice or 6-8 GW in humans, an event mostly completed by GW11 (Fig. 1A).

**Figure 1.**
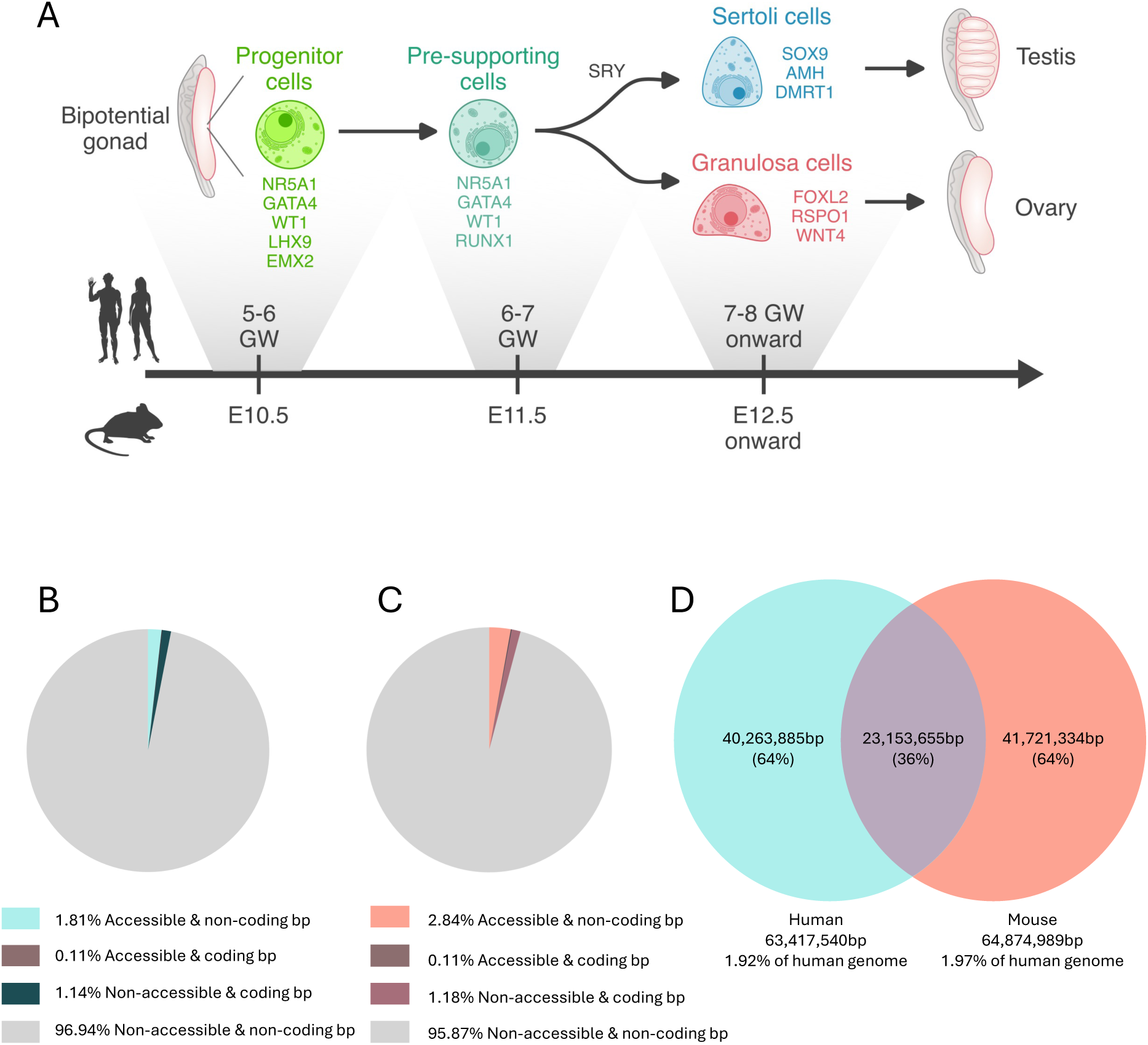
Putative regulatory elements involved in human and mouse gonadal sex determination. (A) Schematic representation of mouse and human gonadal development where early progenitor cells differentiate to pre-supporting cells which later become Sertoli or granulosa in the testis and ovary, respectively. Relevant developmental time points in human and mouse are depicted below. GW- Gestational weeks; E- Embryonic day. (B) Percentage of base pairs (bp) in the human genome (hg38) accessible during human sex determination in pre-supporting cells as well as Sertoli and granulosa cells based on ATAC-seq data, intersected with coding bp. 96.94% of the bp are non-accessible and non-coding. (C) Percentage of bp in the mouse genome (mm10) which are accessible during mouse sex determination in pre-supporting cells as well as Sertoli and granulosa cells based on ATAC-seq data, intersected with coding bp. 95.87% of the bp are non-accessible and non-coding. (D) Intersection of conserved accessible bp in the human and mouse genomes (converted to hg38) during gonadal development in pre-supporting cells as well as Sertoli and granulosa cells based on ATAC-seq datasets. Venn diagram illustrates shared and species-specific accessible bp in human and mouse gonads. Percentages are calculated relative to the human reference genome hg38. bp, base pairs.

To identify the regulatory elements that control murine sex determination, we previously performed ATAC-seq on purified Sertoli and granulosa cells from E11.5, E12.5, E13.5 and E15.5 gonads, spanning the critical developmental window in which early gonad development occurs ^41^. In parallel, to identify regulatory elements involved in early human gonad development, we used publicly available single cell ATAC-seq (scATAC-seq) data from human embryonic gonads between GW6-21 ^37^. To compare the regulatory elements at play during mouse and human gonad development and generate a conserved regulatory atlas, we extracted the open chromatin regions from the cells corresponding to the supporting cell precursors in human XY and XX gonads (termed Pre-Sertoli (PS) or Pre-granulosa (PG)) and from cells corresponding to Sertoli and granulosa cells (Sertoli (SE) or granulosa (GR)) (Supp Fig. 1A, Supp Data 1). This analysis identified 42,026 open chromatin regions corresponding to 63,417,540 nucleotides in human supporting cells, Sertoli and granulosa cells, and representing 1.92% of the human hg38 genome (median peak length of 1,427 bp) (Fig. 1B, Supp Data 1). Just 5.5% of these accessible regions overlap with coding regions. A similar analysis of mouse regulatory elements found 107,837 open chromatin regions corresponding to 83,219,916 nucleotides in supporting, Sertoli and granulosa cells combined, which represent 2.95% of the mouse mm10 genome (median peak length of 678 bp) (Fig. 1C, Supp Data 2). 3.9% of these regions overlap with coding regions of the genome. These data represent a highly relevant set of open chromatin regions, accessible in supporting cell precursors, Sertoli and granulosa cells, in the critical developmental window of early gonad differentiation and development. By contrast, public databases such as ENCODE ^43^ carry data derived from whole gonads, often relying on histone marks, and frequently representing adult or inappropriate foetal stages that are not useful in this context.

To integrate these datasets and construct the first conserved regulatory genome of mouse and human developing gonads, we performed a genome lift of the mouse regulatory elements (mm10) to the human genome (hg38) (see methods). Approximately 75% of the open chromatin peaks identified in mouse were successfully mapped to the human genome (Supp Data 3) and following conversion, they corresponded to 81,324 peaks with 64,874,989 nucleotides, representing ∼2% of the human hg38 genome (median peak length of 672 bp) (Supp Data 4). We intersected this converted mouse data with the open chromatin regions from human embryonic gonad data. This analysis revealed that 36% of the regions accessible in human supporting cells are also accessible in mice, suggesting partial conservation in the regulatory networks in these somatic gonadal cells (Fig 1D, Supp Data 4).

Taken together these analyses show that ∼2-3% of the human or mouse genome, respectively, are putative regulatory elements poised or active in developing gonads. A substantial portion of these regions are conserved between human and mouse. This conserved regulatory atlas provides us with a valuable model system with which to find enhancers instructive of gonad development across these species and to identify and validate variants that may alter regulatory region function in individuals with DSD.

### Identification of a conserved Wt1 enhancer critical for gonadal development in mice

Using the conserved regulatory atlas, we focused on regions likely to function as enhancers and associated with critical genes required for gonad development. One such gene is *Wilms’ tumor 1* (*Wt1*/*WT1*), which encodes a critical transcription factor ^44–46^. Our analysis identified a conserved 984 bp long (mm10, chr2:105,073,954-105,074,937) putative enhancer located upstream of the *Wt1/WT1* gene (Fig 2A, Supp Fig. 2A).

**Figure 2.**
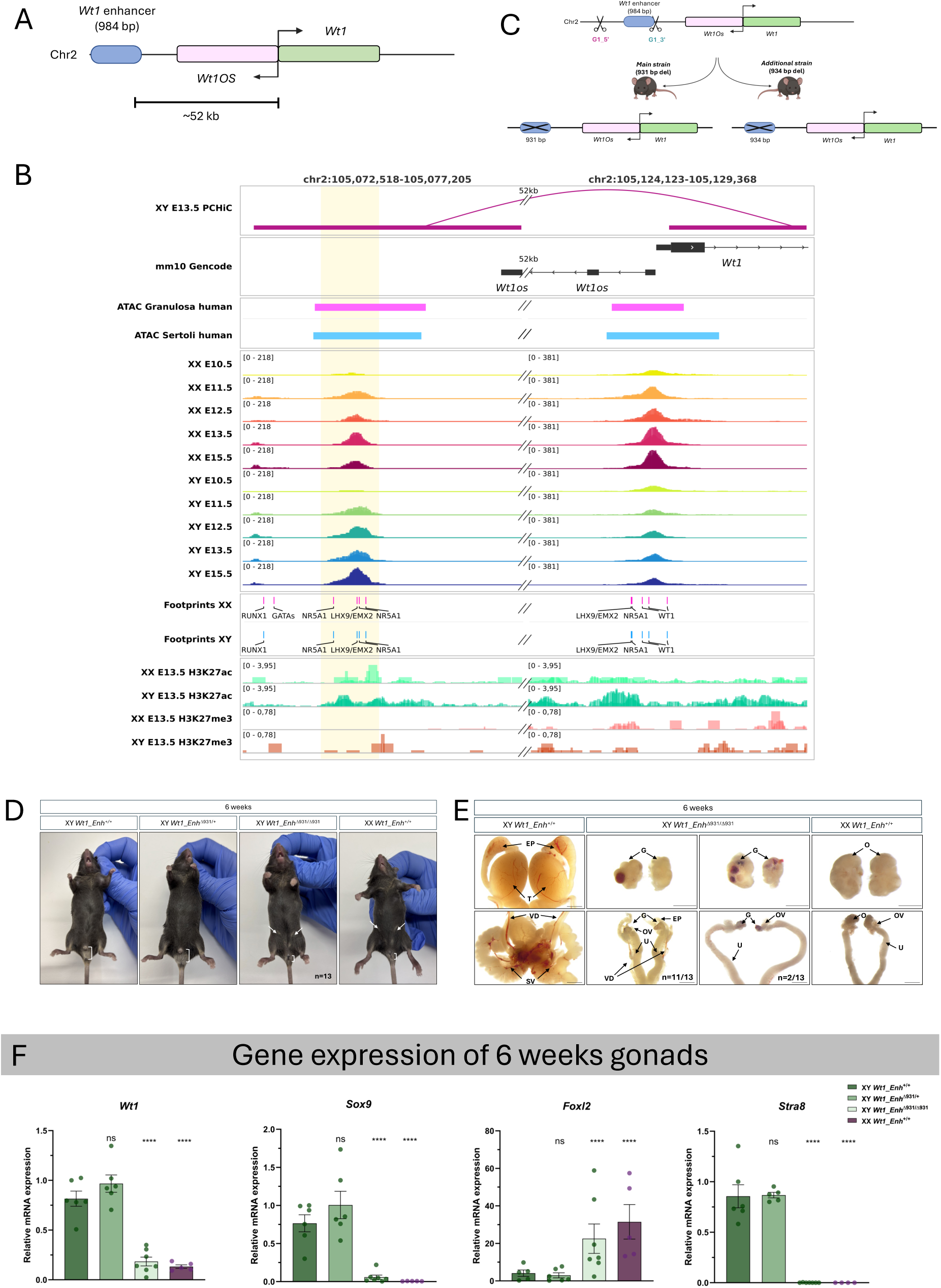
XY mice with feminized external genitalia and dual reproductive tracts upon homozygous deletion of the upstream conserved *Wt1* enhancer. (A) Schematic illustration of the *Wt1* genomic locus containing the long non-coding RNA *Wt1OS* and the upstream enhancer (*Wt1_Enh*) located 52 kb upstream of the *Wt1* transcription start site. (B) Chromatin landscape of the *Wt1*_*Enh* and *Wt1* gene transcription start site region. Tracks shown are the mouse ATAC-seq accessibility of purified supporting cell precursors as well as Sertoli and granulosa cells (E10.5-E15.5), human scATAC-seq accessibility of Sertoli and granulosa cells, transcription factor footprinting, enrichment of active (H3K27Ac) and repressed (H3K27me3) histone marks, and E13.5 promoter capture Hi-C (PCHi-C) interactions linking the upstream enhancer to the *Wt1* promoter. E- Embryonic day (C) Schematic representation of two independent *Wt1_Enh* deletion mouse lines generated by CRISPR-mediated genome editing. (D–E) Representative images of the external genitalia (D) and Bright field images of internal reproductive tracts (E) of 6 weeks-old XY *Wt1_Enh^+/+^*, *Wt1*_*Enh*^Δ931/+^, *Wt1*_*Enh*^Δ931/Δ931^ and XX *Wt1_Enh^+/+^*mice. Brackets indicate anogenital (AG) distance; white arrows indicate nipples. n= number of mice analysed presenting this phenotype. Abbreviations: T, testis; EP, epididymis; VD, vas deferens; SV, seminal vesicles; O, ovary; OV, oviduct; U, uterus; G, gonad. Scale bars represent 2000 µm (testis and reproductive tract) and 1000 µm (gonads). (F) Real-time quantitative PCR analysis of genes involved in gonad differentiation and function in 6 weeks-old gonads of mice carrying the *Wt1_Enh* deletion. Analysis was done for the *Wt1* gene, Sertoli cell marker (*Sox9*), granulosa cell marker (*Foxl2*) and meiosis entry marker (*Stra8*) in XY *Wt1_Enh^+/+^*, *Wt1*_*Enh*^Δ931/+^, *Wt1*_*Enh*^Δ931/Δ931^ and XX *Wt1_Enh^+/+^* gonads. Data are presented as mean 2^−ΔΔCt^ ± SEM and were normalized to the housekeeping gene *Hprt*. Sample size is indicated as dots representing the number of individual mice. Statistical significance was determined using one-way ANOVA followed by Dunnett’s post hoc test. *\*P < 0.05, **P < 0.01, ***P < 0.001,* and *\*\*\*\*P < 0.0001,* ns- not significant. P values below 0.05 were considered statistically significant. All samples are compared to the XY *Wt1_Enh^+/+^*.

The mammalian *Wt1* gene is complex, containing 10 exons with at least 36 reported isoforms created via alternative transcription start site, splicing, and RNA editing ^44,47^. In the mouse, *Wt1* expression starts in the coelomic epithelium and intermediate mesoderm at E9 and continues in the gonads and other tissues ^44^. XY and XX knockout (KO) mice lack gonads and kidneys and die at E13.5 with cardiac and other defects ^44–46^. In humans, pathogenic *WT1* variants are associated with several conditions that include 46, XY DSD as part of the phenotypic spectrum ^44,48^. This includes Wilms’ tumor (OMIM 194070), WAGR syndrome (OMIM 194072), Frasier syndrome (OMIM 136680), Denys-Drash syndrome (OMIM 194080) and Meacham syndrome (OMIM 608978) ^48–51^. Additionally, we previously demonstrated that variants in the 4^th^ zinc finger of *WT1* are a major cause of 46, XX DSD ^52^, consistent with recent demonstration that the *Wt1* (–KTS) splice isoform is essential for ovarian development in mice ^53^.

The putative *Wt1* enhancer identified in our conserved atlas is located ∼52 kb upstream of the mouse *Wt1* transcription start site (Fig. 2A). In mice, this enhancer is not accessible at E10.5 in XY or XX supporting cell precursors but becomes accessible in E11.5–E15.5 Sertoli and granulosa cells ^38,39,41^ (Fig. 2B). It has an active H3K27Ac histone mark in Sertoli and granulosa cells at E13.5 ^38,39,41^ (Fig. 2B). Footprint analysis of transcription factor binding sites (TFBS) in the murine enhancer show consensus sites for NR5A1 and EMX2/LHX9 TFs ^41^ (Fig. 2B). Promoter capture HiC (PCHi-C) assays performed on E13.5 mouse embryonic gonads indicate physical proximity of the *Wt1* promoter with this region in Sertoli cells but not granulosa cells ^41^ (Fig. 2B). Interestingly, this genomic interval has been previously suggested to function as a *Wt1* enhancer exhibiting LacZ reporter activity in Sertoli cells ^40^. The homologous human enhancer is not accessible in XY and XX pre-supporting cell precursors but become accessible in differentiating Sertoli and granulosa cells ^37^ (Fig. 2B).

We used CRISPR genome editing to delete this enhancer in mice (Fig. 2C). Two independent mouse strains were generated containing a deletion of this enhancer (Fig. 2C, Supp Fig. 2B-C, a 931 bp deletion (*Wt1*_*Enh*^Δ931^) and a 934 bp deletion (*Wt1*_*Enh*^Δ934^)). Founder mice carrying these deletions were crossed with *wild type* mice for germline transmission, and two heterozygous littermates were subsequently inter-crossed to generate homozygous animals for analysis. Analysis of the first mouse line with a 931 bp deletion, removing most of the *Wt1* enhancer (Fig. 2C, Supp Fig. 2B), demonstrated that while adult XY *Wt1*_*Enh*^Δ931/+^ mice developed as phenotypic males and were fertile, all adult XY *Wt1*_*Enh*^Δ931/Δ931^ mice were phenotypic females externally – presenting with short anogenital distance and nipples, similar to XX *Wt1*_*Enh*^+/+^ females (n=13) (Fig 2D). Analysis of the internal reproductive organs revealed that 15% of XY *Wt1*_*Enh*^Δ931/Δ931^ mice (n=2/13) had a fully feminized reproductive system albeit with dysgenic ovarian-like gonads as well as an oviduct, uterus and vagina. However, the majority exhibited persistent Müllerian and Wolffian derivatives, with dysgenic ovarian-like gonads, oviducts, a uterus and small epididymides and vas deferens (n=11/13). The gonads of XY *Wt1*_*Enh*^Δ931/Δ931^ mice were smaller than ovaries of XX *Wt1*_*Enh*^+/+^ counterparts and most had haemorrhagic lesions (Fig 2E, Supp Fig 3A).

**Figure 3.**
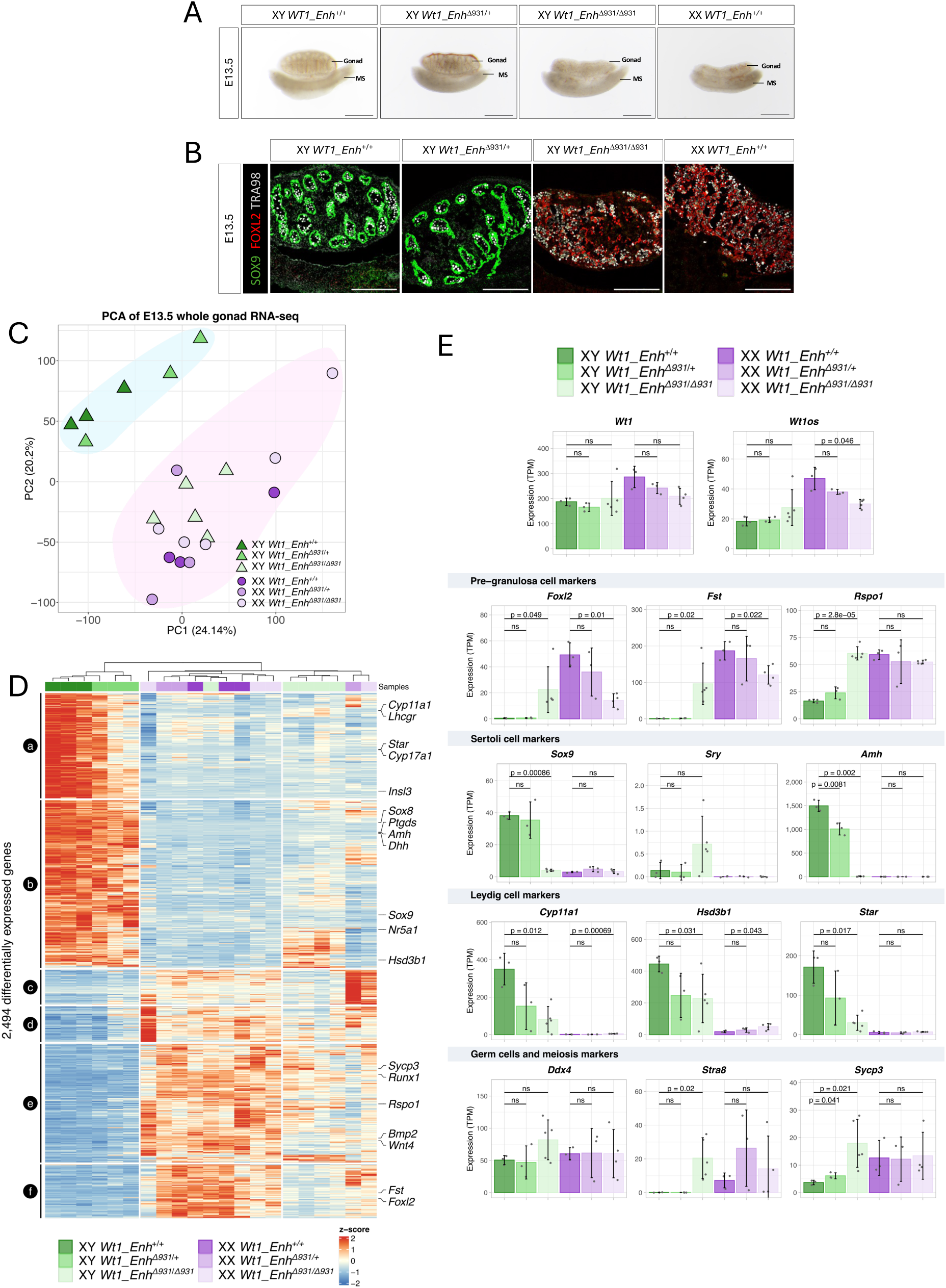
XY male-to-female sex reversal in homozygous embryos lacking the upstream conserved *Wt1* enhancer. (A) Brightfield images of E13.5 gonads from XY *Wt1_Enh^+/+^*, *Wt1*_*Enh*^Δ931/+^, *Wt1*_*Enh*^Δ931/Δ931^ and XX *Wt1_Enh^+/+^*embryos. MS-mesonephros, E-Embryonic day. Scale Bars represent 500 µm. (B) Immunostaining of E13.5 gonads from XY *Wt1_Enh^+/+^*, *Wt1*_*Enh*^Δ931/+^, *Wt1*_*Enh*^Δ931/Δ931^ and XX *Wt1_Enh^+/+^*embryos. Gonads were stained for Sertoli-marker SOX9 (green), granulosa-marker FOXL2 (red) and germ cells marker TRA98 (grey). Scale bars represent 200 µm. (C) PCA of the transcriptomes from E13.5 XY and XX whole gonads of *Wt1_Enh^+/+^*, *Wt1*_*Enh*^Δ931/+^ and *Wt1*_*Enh*^Δ931/Δ931^ embryos. Each point represents the transcriptome of a pair of gonads, separated from the mesonephros. Samples are coloured by their genotype, and the shape of the point represents the sex of the gonads (circle- XX, triangle- XY). (D) Heatmap representing the differentially expressed genes between the samples. Expression data were normalized as Z-scores. Differentially expressed genes were clustered into 6 groups labelled from *a* to *f* as indicated on the left-hand side of the heatmap. Samples were grouped using unsupervised clustering as represented by the dendrogram on the top of the heatmap. Names of well-known gonadal marker genes are indicated on the left-hand side. (E) Expression profiles of well-known gonadal marker genes including *Wt1* and *Wt1os* as well as markers of Sertoli cells, pre-granulosa cells, Leydig cells and germ cells & meiosis cell markers. Expression values are expressed as TPM (Transcript per million), the bars represent the median of expression, each point represent a single biological replicate. Error bars represent the standard deviation across the replicates. Statistical significance was determined using one-way ANOVA followed by Dunnett’s post hoc test.

Immunostaining of gonads from mice at 6 weeks was performed using antibodies for SOX9 to mark Sertoli cells, FOXL2 to mark granulosa cells, and TRA98 to mark germ cells (Supp Fig 3B-C). XY *Wt1*_*Enh*^+/+^ and *Wt1*_*Enh*^Δ931/+^ mice had gonads with seminiferous tubules containing SOX9-expressing Sertoli cells and spermatids marked by TRA98. XX *Wt1*_*Enh*^+/+^ mice had gonads with follicles containing FOXL2-expressing granulosa cells engulfing oocytes marked by TRA98. XY *Wt1*_*Enh*^Δ931/Δ931^ mice had gonads with an ovarian-like morphology with follicle structures, FOXL2, and mostly devoid of SOX9 expression (Supp Fig 3B). We carried out further analysis of the WT1 protein as well as Steroidogenic Factor 1 (SF1, encoded by *Nr5a1*), a known target of WT1 in the gonads ^54,55^. Immunostaining with in XY gonads from *Wt1*_*Enh*^+/+^ and *Wt1*_*Enh*^Δ931/+^ mice demonstrated WT1 and SF1 expression in Sertoli cells within seminiferous tubules and strong SF1 expression within Leydig cells located in the interstitial compartment. *Wt1*_*Enh*^+/+^ XX mice had WT1 and SF1 expression in granulosa cells within follicles whereas SF1 is strongly expressed in theca and steroidogenic cells of the ovary. XY *Wt1*_*Enh*^Δ931/Δ931^ mice still had some WT1 protein present, as well as SF1 in a highly disorganised ovarian-like gonad (Supp Fig 3C).

We then validated the phenotype in a second, independent mouse line carrying 934 bp deletion (*Wt1*_*Enh*^Δ934^), eliminating most of the *Wt1* enhancer (Fig. 2C, Supp Fig. 2C). Consistent with the first mouse line, XY *Wt1*_*Enh*^Δ934/Δ934^ mice also exhibited male-to-female sex reversal and presented with feminised phenotype characterized by short anogenital distance and nipples. Internally, they displayed dysgenic gonads with reproductive tracts containing both Müllerian derivatives (uterus and oviducts) and Wolffian derivatives (epididymis and vas deferens) (Supp Fig. 3D). In summary, bi-allelic deletion of a *Wt1* enhancer identified in a conserved regulatory atlas caused XY sex reversal in mice.

We next assessed whether deletion of the *Wt1* enhancer also affected XX gonadal development. *Wt1*_*Enh*^+/+^, *Wt1*_*Enh*^Δ931/+^ and *Wt1*_*Enh*^Δ931/Δ931^ XX adults all presented as phenotypic females with short anogenital distance and nipples (Supp Fig. 4A). Internally, ovaries, oviduct and uterus were present in all lines. Closer analysis of *Wt1*_*Enh*^Δ931/Δ931^ XX female ovaries revealed they were smaller compared to *Wt1*_*Enh*^+/+^ and *Wt1*_*Enh*^Δ931/+^ and often presented with red spots likely to be haemorrhagic lesions (Supp Fig. 4A-B). Quantification of ovary weight in 6-week-old adults revealed that *Wt1*_*Enh*^Δ931/+^ females had comparable ovary/kidney or ovary/body weight ratios to that of *Wt1*_*Enh*^+/+^ females yet *Wt1*_*Enh*^Δ931/Δ931^ females presented with ∼45% decrease in relative ovary weight (Supp Fig. 4C-D). No change in kidney weight or in overall morphology was noted for any mice (Supp Fig. 4E), suggesting the importance of this enhancer in driving *Wt1* expression might be restricted to gonad development. The decrease in ovary size and haemorrhagic lesions was also observed in the second independent mouse line carrying a 934 bp deletion (Supp Fig. 3D). We further analysed reproductive capacity. Fertility tests, in which adult XX *Wt1*_*Enh*^+/+^ and *Wt1*_*Enh*^Δ931/Δ931^ females were bred with *Wt1*_*Enh*^+/+^ males for a period of six months revealed that *Wt1*_*Enh*^+/+^ females had an average of ∼ 4 litters during a six month period while XX *Wt1*_*Enh*^Δ931/Δ931^ females only had 1 litter or no litters at all, suggesting a major sub-fertility in XX *Wt1*_*Enh*^Δ931/Δ931^ females (Supp Fig. 4I). The average number of pups born per litter did not show any significant change (Supp Fig. 4I).

**Figure 4.**
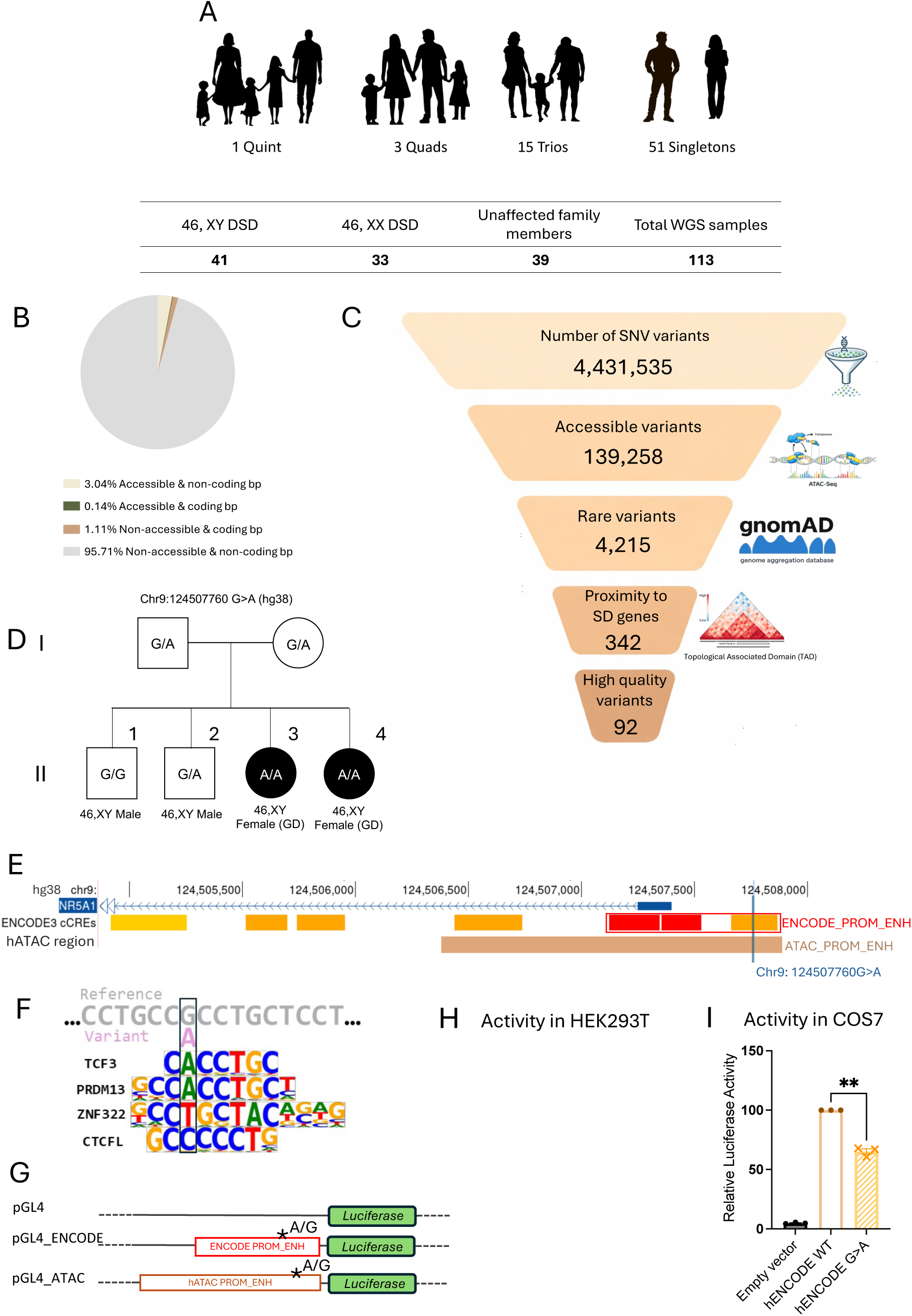
Multi-modal filtration pipeline identifies novel variant in conserved enhancer upstream of *NR5A1*. (A) Composition of the DSD cohort analyzed in this study. Short-read whole genome sequencing (WGS) was performed on individuals with DSD and their family members. (B) Percentage of base pairs (bp) in the human genome (hg38) which are accessible during either human or mouse sex determination based on ATAC-seq datasets, intersected with coding bp. 95.71% of the bp are non-accessible and non-coding. (C) Multi-modal variant prioritization pipeline used to identify variants in DSD patients. Single-nucleotide variants (SNVs) identified by whole-genome sequencing were filtered to retain variants located within either human-specific, mouse-specific or human-mouse conserved chromatin-accessible regions identified in embryonic gonads. Rare variants were then retained based on population allele frequency (gnomAD), followed by prioritization of variants located near sex-determination (SD) genes within the same topologically associating domain (TAD), or by genomic distance when TAD information was not available. Application of quality filters retained a set of high-confidence candidate variants. Numbers represent the average number of SNVs per sample after each filtering step from a cohort of 74 DSD patients. (D) Family pedigree for variant upstream of *NR5A1*. The G>A change (Chr9:124,507,760), inherited from heterozygous parents, is homozygous in the affected 46,XY female children (II.3 and II.4), and heterozygous or *wild type* in the unaffected 46,XY siblings. (E) The variant (blue arrow and line) lies within an open chromatin region in human and mouse ATAC-seq datasets (black line) and within a predicted proximal enhancer (ENCODE). (F) Luciferase vectors cloned and tested. (G-H) Relative luciferase activity of the ENCODE promoter-enhancer regions with and without the patient variant in HEK293T, p-value 0.0005 (G) or COS7 cells p-value 0.0041 (H). The variant causes a significant reduction in activity. Each data point represents the mean from three technical replicates, with three biological replicates (independent experiments) shown. Bars are SEM. P-values calculated with unpaired student T-test.

Immunostaining of ovaries from 6-week-old XX mice showed FOXL2 expression in *Wt1*_*Enh*^+/+^*, Wt1*_*Enh*^Δ931/+^ and *Wt1*_*Enh*^Δ931/Δ931^ mice, who were all similarly devoid of SOX9 expression. TRA98 positive oocytes were detected in some follicles. While *Wt1*_*Enh*^Δ931/Δ931^ XX mice expressed FOXL2 in their ovaries, their follicles appeared less organised compared to controls (Supp Fig. 4G). WT1 protein was detected in granulosa and stromal cells, while SF1 expression was observed in granulosa, theca and steroidogenic cells of the ovary (Supp Fig. 4H). In summary, loss of both copies of the *Wt1* enhancer in XX mice resulted in a reduction in ovarian weight, subtle structure disorganisation, and significant subfertility.

To further explore the gonadal expression profiles of adult XY and XX *Wt1*_*Enh*^Δ931/Δ931^ mice, we performed qPCR analysis on selected gonadal markers (Fig. 2F, Supp Fig. 5A-B). Testis from XY *Wt1*_*Enh*^Δ931/Δ931^ mice had a 77% decrease in *Wt1* expression levels compared to *Wt1*_*Enh*^+/+^ testes, resulting in levels comparable to that of *Wt1*_*Enh*^+/+^ XX ovaries (Fig. 2F). *Sox9* and *Sox8* expression levels were also reduced by more than 90% and were comparable to *Wt1*_*Enh*^+/+^ XX ovaries, further supporting the XY sex reversal phenotype (Fig. 2F, Supp Fig. 5A). Supporting this, granulosa markers including *Foxl2* and *Rspo1* were significantly elevated in XY *Wt1*_*Enh*^Δ931/Δ931^ mice, reaching levels similar to those observed in XX *Wt1*_*Enh*^+/+^ gonads (Fig. 2F, Supp Fig. 5A). No significant change was observed in the expression levels of *Nr5a1* and *Gata4.* Strikingly, while XY *Wt1*_*Enh*^+/+^ and *Wt1*_*Enh*^Δ931/+^ mice displayed high expression levels of *Stra8*, a meiosis entry marker, resembling active spermatogenesis in adult testis, XY *Wt1*_*Enh*^Δ931/Δ931^ mice had no *Stra8* expression, similarly to XX *Wt1*_*Enh*^+/+^ ovaries (Fig. 2F). *3ßHsd,* an embryonic Leydig cell marker, had a remarkable 50-fold elevation in expression in XY *Wt1*_*Enh*^Δ931/Δ931^ mice compared to XY *Wt1*_*Enh*^+/+^ (Supp Fig. 5A). Lastly, no major change in the expression levels of the steroidogenic-precursor marker *Nr2f2* was seen (Supp Fig. 5A).

XX *Wt1*_*Enh*^Δ931/Δ931^ ovaries displayed 40% decrease in *Wt1* expression levels but had no significant change in the expression levels of *Nr5a1*, *Gata4*, *Stra8* and *3ßHsd.* A small, yet statistically significant increase was seen in the levels of *Sox9* and *Sox8* in XX *Wt1*_*Enh*^Δ931/Δ931^ compared to XX *Wt1*_*Enh*^+/+^. No significant change was seen in the expression levels of *Foxl2*, yet a 55% decrease in the expression levels of *Rspo1* and ∼3 fold increase in the expression levels of *Nr2f2* were noted (Supp Fig. 5B).

Taken together, these data suggest this conserved upstream enhancer of *Wt1* is critical for proper testis and ovary development and function in mice. In its absence, XY mice present male-to-female sex reversal whereas XX mice display ovarian disorganisation and sub-fertility.

### The Wt1 enhancer is critical at embryonic timepoints during gonadal development

To further investigate the function of this enhancer in regulating *Wt1* expression during embryonic gonad development, we analysed E13.5 mouse gonads from all genotypes. XY *Wt1*_*Enh*^+/+^ and *Wt1*_*Enh*^Δ931/+^ gonads exhibited testis cords and a coelomic vessel, characteristic of embryonic testis. XY *Wt1*_*Enh*^Δ931/Δ931^ gonads presented an ovarian morphology, lacking testis cords and a defined coelomic vessel (Fig 3A). XX *Wt1*_*Enh*^+/+^, *Wt1*_*Enh*^Δ931/+^ and *Wt1*_*Enh*^Δ931/Δ931^ embryos all presented with typical ovarian morphology (Supp Fig 6A). Immunostaining of E13.5 gonads (Fig 3B) (Supp Fig 6B) found that XY *Wt1*_*Enh*^+/+^ and *Wt1*_*Enh*^Δ931/+^ gonads had SOX9^+^, GATA4^+^, WT1^+^ Sertoli cells, surrounding the TRA98^+^ gonocytes, with SF1 expression detected in both Sertoli cells and Leydig cells residing in the interstitial compartment. WT1 was also expressed in the mesonephros. XY *Wt1*_*Enh*^Δ931/Δ931^ embryos had gonads lacking SOX9 expression but showed FOXL2 and TRA98 expression in ovarian-like structures (Fig 3B). GATA4, WT1 and SF1 expression was also detected in a pattern similar to the ovarian morphology in XX *Wt1*_*Enh*^+/+^ gonads (Supp Fig 6B). WT1 expression was also evident in the mesonephros of XY *Wt1*_*Enh*^Δ931/Δ931^ embryos. Similar analysis on XX *Wt1*_*Enh*^+/+^, *Wt1*_*Enh*^Δ931/+^ and *Wt1*_*Enh*^Δ931/Δ931^ revealed a normal ovarian expression pattern (Supp Fig 6C-D).

RNA-seq was performed on pairs of E13.5 gonads from XY and XX of *Wt1*_*Enh*^+/+^, *Wt1*_*Enh*^Δ931/+^ and *Wt1*_*Enh*^Δ931/Δ931^ embryos (Fig 3C-E, Supp Fig 7). Principal component analysis (PCA) demonstrated that XY *Wt1*_*Enh*^+/+^ and *Wt1*_*Enh*^Δ931/+^ gonads clustered together, and all XX gonads clustered together, suggesting that the enhancer deletion does not significantly impact embryonic ovary identity in an XX background. On the contrary, gonads from XY *Wt1*_*Enh*^Δ931/Δ931^ mice clustered with XX ovaries, suggesting the complete XY male-to-female sex reversal phenotype is also prevalent at the transcriptomic level (Fig 3C). Spearman pairwise sample correlation analysis supported these findings and separated the XY *Wt1*_*Enh*^+/+^ and *Wt1*_*Enh*^Δ931/+^ samples from both the XX samples and XY *Wt1*_*Enh*^Δ931/Δ931^ gonads (Supp Fig 7A).

Differential expression analysis identified 2,494 genes differentially expressed, classified into 6 groups (*a* to *f*) based on their expression profile (Fig 3D, Supp Data 5). Groups *a*-*b* exhibited increased expression in the testis while groups *c*-*f* constitute genes with increased expression in the ovary. Consistent with the sex reversal phenotype, XY *Wt1*_*Enh*^Δ931/Δ931^ gonads had a transcriptomic profile resembling ovaries rather than testis (Fig 3D). This includes elevated expression of ovarian markers, decreased Sertoli cell marker expression and elevated expression of Leydig markers (present at the bottom of cluster *b*) that are normally only expressed in XY gonads. This expression of Leydig cell markers aligns with the presence of Wolffian duct derivates in XY *Wt1*_*Enh*^Δ931/Δ931^ mice giving rise to the epididymis and vas deference in response to androgen production (Fig. 2E). Interestingly, while four XY *Wt1*_*Enh*^Δ931/Δ931^ gonads clustered together, one XY *Wt1*_*Enh*^Δ931/Δ931^ gonad clustered more closely to XX ovaries – potentially representing a mouse with the less abundant phenotype of female-only reproductive tracts (Fig. 2E).

This RNA-seq data revealed that *Wt1* and its non-coding partner, *Wt1os,* do not show a significant decrease in expression at these stages, in contrast to our findings in adult stages (Fig 3E). Similarly, *Gata4*, which is expressed in supporting cell precursors, and later maintained in both Sertoli and pre-granulosa cells, also revealed no major differences across phenotypes (Supp Fig 7B). In contrast *Nr5a1* (SF1) expression was significant reduced in both XY *Wt1*_*Enh*^Δ931/+^ and *Wt1*_*Enh*^Δ931/Δ931^ gonads compared to XY *Wt1*_*Enh*^+/+^gonads (Fig 3E). *Sox9*, *Sox8* and *Fgf9* also showed a significant decrease in XY *Wt1*_*Enh*^Δ931/Δ931^ gonads, consistent with the absence of Sertoli cells (Fig 3E, Supp Fig 7B). Likewise, *Amh* expression was decreased, explaining the persistence of Müllerian duct derivatives such as oviduct and uterus in XY *Wt1*_*Enh*^Δ931/Δ931^ mice ^56^ (Fig 3E). *Desert Hedgehog* (*Dhh*), normally secreted from Sertoli cells and important for Leydig cell specification ^57^, was also downregulated in XY *Wt1*_*Enh*^Δ931/Δ931^ gonads, raising the question of how Leydig cells are being specified in this context (Supp Fig 7B). Interestingly, *Sry* transcripts were elevated in XY *Wt1*_*Enh*^Δ931/Δ931^ gonads. *Sry* expression is known to be repressed upon induction of SOX9 in Sertoli cells ^58^, meaning this observation supports the inability to initiate the testicular differentiation and *Sox9* activation (Fig 3E). Analysis of pre-granulosa markers showed a reciprocal pattern with strong upregulation in the expression of *Foxl2, Fst*, *Rspo1*, *Bmp2*, *Wnt4*, *Runx1* (Fig 3E, Supp Fig 7B). For some markers the expression levels in XY *Wt1*_*Enh*^Δ931/Δ931^ gonads were comparable to XX *Wt1*_*Enh*^+/+^, while for other genes, the expression level did not reach the XX *Wt1*_*Enh*^+/+^ but was markedly higher than that in XY *Wt1*_*Enh*^+/+^ and *Wt1*_*Enh*^Δ931/+^ gonads. Analysis of multiple Leydig markers including *Cyp11a1, Hsd3b1*, *Star*, *Insl3*, *Cyp17a1* and *Lhcgr* revealed that while XY *Wt1*_*Enh*^Δ931/Δ931^ gonads have a significant decrease in expression, it is not fully abolished as in the XX gonads. (Fig 3E, Supp Fig 7B). Finally, germ cell markers including *Ddx4*, *Dazl*, and *Nanos3* were largely unchanged. Strikingly, the expression of *Stra8*, a meiosis entry hallmark gene ^59^, and *Sycp3*, suggested germ cell entry to meiosis in XY *Wt1*_*Enh*^Δ931/Δ931^ gonads, similar to the XX gonads (Fig 3E, Supp Fig 7B).

Taken together, these data suggest that complete deletion of this *Wt1* enhancer causes XY male-to-female sex reversal from early embryonic stages where XY homozygous gonads fail to give rise to Sertoli cells and instead adopt a pre-granulosa cell fate. Surprisingly, in most cases, Leydig cells are also present in these XY homozygous gonads, supporting androgen synthesis and resulting in the presence of both Müllerian duct and Wolffian duct derivatives in the same animal. In XX embryos, the deletion of this enhancer element results in an ovarian phenotype, which only becomes apparent in adults, suggesting distinct, sex-specific roles for the gonadal enhancer during gonad development and maturation.

### Variants in the WT1 enhancer may underlie 46,XX and XY DSD

Given the strong gonadal phenotype resulting from deletion of the murine upstream *Wt1* enhancer, we wanted to further explore this enhancer in the human context. In humans this enhancer is both conserved and accessible in embryonic gonads (Fig. 2B, Supp Fig 2A). Hence, we analysed this region in a cohort of >400 undiagnosed DSD patients using either WGS data or Sanger sequencing targeted to this region. This analysis identified a total of 8 patients carrying rare variants within this enhancer region (Supp Fig 8A). Interestingly, Patient No. 1 who presented with 46,XY gonadal dysgenesis had a left streak gonad with Müllerian remnant and a right testis with epididymis of normal size and vasculature – similar to the phenotype observed in mice. *In silico* analysis demonstrates that several of these variants lie within putative transcription factor binding sites (TFBS) (Supp Fig 8B). Further, an analysis of genetic constraint of this *WT1* enhancer region revealed a weighted Z score of 1.55, which exceeds both the median Z score of 0.08 for non-coding sequences and 1.48 for all protein-coding sequences in the human genome. Thus, variants in this enhancer region may contribute to DSD.

### A computational filtration pipeline refines potential causative variants in WGS of DSD individuals

The above results have demonstrated the power of a conserved gonadal regulatory atlas in identifying *c*REs essential for gonad development in mice and that may be associated with DSD in humans. To further extend this analysis, we used this conserved regulatory atlas to develop a multi-modal computational filtration pipeline for variants in non-coding regions from undiagnosed DSD with disrupted gonadal development. This integrated the conserved accessible open chromatin regions from mouse and human embryonic gonads, retaining the human-specific intervals, the mouse-specific intervals converted to human genome, and the mouse & human conserved enhancers (in total representing 3.18% of the human hg38 genome) (Fig 4B, Supp data 4). The decision to retain mouse-specific intervals, converted to the human genome, is because our pipeline uses human single cell ATAC-seq data meaning that these important regions may have been missed due to the relatively shallow sequencing depth when compared to bulk ATAC-seq on purified relevant cell types. We then collated WGS data from a cohort of individuals with DSD who remained without a genetic diagnosis following exome or gene panel sequencing. This cohort includes a total of 74 DSD individuals; 41 with 46,XY DSD (ranging from under-virilisation, partial gonadal dysgenesis to complete gonadal dysgenesis or ovo-testicular DSD) and 33 with 46,XX DSD (including ovo-testicular DSD and ovarian dysgenesis). 51 individuals were singletons and the rest were sequenced as a trio or a larger family (Fig 4A, Supp data 6). All patients were sequenced using the Illumina or BGI short read WGS platforms and we analysed small indels or single nucleotide variants (SNV) from these patients.

WGS of the 74 individuals with DSD identified, on average, 4,431,535 small indels or SNVs per patient (Fig 4C). Our pipeline filtered these variants to retain those located within a conserved putative regulatory region defined in our atlas (Supp data 4, Supp data 6). This eliminated ∼97% of the variants, leaving an average of 139,258 variants per patient (Fig 4C, Supp data 6). We then filtered for frequency, using data from the Genome Aggregation Database (GnomAD V4.0), composed of 76,215 WGS datasets. Variants with a population maximum (popmax) allele frequency greater than 0.001 were excluded, reducing the number to an average of 4,215 variants per individual and removing 99.9% of the initial variants (Fig 4C, Supp data 6). Next, we curated a list of 105 genes known or suspected to play a critical role in gonadal development or DSD based on human and mouse studies (Supp data 7). We extracted Topologically Associated Domains (TADs) for these genes from human embryonic stem cell Hi-C data ^60^ (Supp data 7) as TADs are considered to be highly stable among different cell types ^61^. For genes lacking TAD information, we used a +/– 1.5 MB window from the gene transcription start site/end site. We then filtered for variants located within TADs of these 105 genes, resulting in an average of 342 variants per patient. Finally, internal quality assessment removed variants located in highly repetitive genomic regions, or which were frequent in our cohort (>30%) representing possible sequencing artifacts (see methods). This left us with an average of 92 variants per individual with some individuals having as few as 8 variants and others 280 variants (Fig 4C, Supp data 6). In summary, this multi-modal filtration pipeline allowed us to refine and prioritise candidate variants from a starting point of 4.4 million variants on average to 92 variants per individual, revealing candidate non-coding variants for DSD to be further tested and functionally validated.

### A homozygous variant in a novel enhancer region of NR5A1 segregates with gonadal dysgenesis in a pedigree

Our DSD cohort contained several trios and larger families, allowing us to further filter variants using familial segregation. To this end, we adapted our filtration pipeline to include familial segregation for the common inheritance modes associated with DSD ^7^. These include *de novo*, homozygous, compound heterozygous and sex-limited inheritance (a variant found in 46,XY DSD inherited from a healthy XX mother or *vice versa*). One family included in our cohort has two unaffected parents, two unaffected 46,XY sons and two 46,XY daughters with gonadal dysgenesis (Supp data 6, Fig 4D). WGS from five family members (the affected individuals, parents, and one unaffected XY sibling) was analysed in our multi-modal filtration pipeline. Using this approach, we identified a homozygous, single nucleotide variant, located 361 bp upstream of the *NR5A1* gene (encoding the Steroidogenic Factor 1 (SF1) protein). This variant (Chr9:124,507,760 G>A, rs958814655, hg38) is homozygous in the two affected siblings, heterozygous in the two unaffected parents (Fig 4D, Supp Fig 9A) and one unaffected 46,XY brother, whereas a second unaffected 46,XY brother was *wild type* for the variant, as confirmed by Sanger sequencing (Fig 4D, Supp Fig 9A).

SF1/*NR5A1* is a known regulator of gonadal development, and variants in this gene are a well-established cause of DSD. The phenotypes observed in the two affected children are consistent with pathogenic variants in *NR5A1* reported in other DSD cases ^62–66^. Specifically, the younger 46,XY sister (II.4, Supp Fig 9A) presented in their teens with virilisation at puberty and a history of clitoromegaly from birth. She was found to have a 46,XY karyotype and was diagnosed with partial gonadal dysgenesis. Histology revealed a right streak gonad with fallopian tube, and a left atrophic testis with spermatic cord and epididymis (Supp Fig 9B-D). Her older sister (II.3) presented with delayed puberty and was also found to have a 46,XY karyotype. Histology revealed small left and right gonadal remnants with ovarian-like stroma and fallopian tubes on both sides. A very small vas-deferens like structure was noted on the right (Supp Fig 9E-G). She had no virilisation and was diagnosed with complete gonadal dysgenesis. Microarray analysis found that both affected sisters shared 3 long continuous stretches of homozygosity (each region >2 Mb) on chromosomes 3 and 9, suggesting consanguinity in the family. The variant falls within one of these regions (chr9:120,976,387-135,190,019). WES analysis had previously excluded any other likely causative variants, including dominant or recessive rare damaging variants that segregated in the family and affecting known DSD genes.

This non-coding region upstream of *NR5A1* variant is reported in two instances as heterozygous in GnomAD v4.1 (one in an admixed American population and the other in “remaining”), with no homozygous variants described in the remaining 152,146 alleles in this version. We also interrogated several other regional genomic databases. In the Hong Kong Genome Project database with 10,000 genomes from predominantly Chinese heritage just one heterozygous individual was identified. In an additional database of 85 genomes of Filipino ancestry (<u>OurDNA Browser</u>), this variant was not found.

The G>A variant falls within a predicted proximal enhancer region from ENCODE (Fig 4E) and is upstream of two predicted promoter regions for the human *NR5A1* gene. The affected nucleotide is conserved in mammals from humans to elephants (Supp Fig 9H) and is accessible in mouse gonads (Supp Fig 9I). Transcription factor binding analysis (using JASPAR 2026 core set in UCSC with a minimum score of 360) at this site in humans revealed that the affected G nucleotide sits within five predicted TFBSs. The first, for CTCFL (reverse orientation), the although the G>A change does not affect this consensus. The second is in a TFBS for PRDM13, and the G>A change creates a preferred site at this nucleotide, (binding score = 12.9 in *wild type*, 17.2 in mutant; FIMO) (Fig 4F). The third is a weakly implicated ZNF322 consensus binding site. Interestingly, by relaxing the minimum score for the WT sequence, we found that the G>A replacement creates a perfect TCF3 (Fig 4F, binding score = 9.43 in *wild type* and 12.86 in mutant, FIMO) and TCF12 (Fig 4F, binding score = 9.93 in *wild type* and 12.61 in mutant, FIMO). This is of interest as TCF3, a basic helix-loop-helix transcription factor (also known as E2A) is a transcriptional repressor that is which has recently been suggested as a novel factor predicted to play a pivotal role in gonadal development ^67^, while TCF12 is highly expressed in supporting somatic cells in embryonic gonads ^68^. For this enhancer, genomic constraint analysis revealed a weighted Z score of 1.36, well above the median score of 0.08 for non-coding sequences and just below the median of 1.48 for protein coding sequences, again suggesting a moderate constraint for this enhancer sequence compared to other non-coding regions.

While the nucleotide is conserved in mouse, there is misalignment of surrounding nucleotides, with the mouse genome harbouring a small deletion of 7 nucleotides adjacent to the affected nucleotide. This means that predicted TFBSs at this site are not present in the mouse. Given it is likely that the activity of this region and variant may differ in mice, we assessed the activity in a human assay. To assess the activity of the predicted human enhancer region and the patient variant in a human cellular context, we cloned two regions from the human genome into a Luciferase reporter vector. The first construct (1.7 kb) encompasses the combined ATAC-seq region from the human gonadal dataset (hATAC), spanning the upstream predicted enhancer from ENCODE3 into the first intron of *NR5A1* (chr9:124506219-124507940, Fig. 4E,G, Supp Fig 10A). A second smaller construct (844 bp) encompasses the ENCODE predicted proximal enhancer-like signature of interest (enhP129, encode accession EH38E2726129) as well as two predicted promoter sequences (chr9:124507097-124507940, encode accession EH38E2726127-128) (Fig 4E,G, Supp Fig 10A). Both regions were cloned into an empty pGL4 luciferase reporter vector after amplification from an unaffected *wild type* brother. Sanger sequencing after cloning confirmed that no polymorphisms were present in this region of DNA compared to the reference genome. Transfection was performed alongside an empty vector control, and with a Renilla transfection control plasmid, into Human Embryonic Kidney 293T cells (HEK293T). Both regions showed considerable activity with the hATAC region, showing around 30-fold induction compared to an empty vector control (Supp Fig 10A). Given that the ENCODE region was smaller and had slightly higher activity, we decided to proceed with this region to test the effect of the variant. We cloned this region from the proband, with Sanger sequencing confirming that the only difference to the unaffected brother was the G>A change of interest at Chr9:124,507,760. When the vector carrying this variant was transfected into HEK293T cells, it showed on average a 30% decrease in activity compared to the *wild type* ENCODE region (p-value 0.0005, n=3) (Fig 4H). To confirm this, we repeated this assay in COS7 cells, where we found a 35% average reduction in activity compared to the *wild type* allele (p-value 0.0041, n=3) (Fig 4I). To further confirm that the predicted proximal enhancer containing the variant contributes to the observed reduction of activity, this enhancer alone (enhP129, encode accession EH38E2726129) was cloned into a luciferase vector upstream of a minimal promoter (Supp Fig 10C-D), both with and without the variant. In HEK293T cells, this enhancer fragment upregulated the activity of the minimal promoter by 4.5-fold on average. This activation was reduced to 2.4-fold when the G>A variant was introduced (46% reduction, p-value 0.0072 n=3) (Supp Fig 10D), demonstrating that this single nucleotide change causes a strong reduction in enhancer activity.

Therefore, using our conserved gonadal regulatory atlas pipeline, we have identified an enhancer for *NR5A1* with *in vitro* activity, in which a bi-allelic single nucleotide variant was found, segregating with 46,XY gonadal dysgenesis in a family. This G>A variant significantly reduces the activity of this enhancer in luciferase assays. TFBS analysis indicates that this variant falls within several predicted TFBSs and creates a perfect binding site for the known transcriptional repressor, TCF3. We postulate that this variant causes a reduction in *NR5A1* during a critical period of gonadal development leading to gonadal dysgenesis.

## Discussion

In this study, we created the first inter-species conserved regulatory atlas for early gonadal development ^37,41^. Our analysis shows that 36% of gonadal putative regulatory elements are conserved between mouse and human somatic cells. These findings are in accordance with a recent comparison of single cell transcriptomics data from the mouse and human embryonic gonads, where transcriptomic differences were found in central gonadal cell types with the fully differentiated cells showing greater similarity between species compared to their progenitor cells ^19^.

### A *Wt1* upstream enhancer is crucial for gonad development

Our atlas identified a conserved and functionally critical enhancer for *Wt1*. *Wt1/WT1* expression is complex and widespread ^44–46^, yet the deletion of this *Wt1* enhancer, identified in our atlas, resulted in a clear gonad-specific phenotype, suggesting its major importance for expression in this tissue. In the gonads, this enhancer seems to have dual, temporally-distinct roles between sexes. In XY gonads, loss of this enhancer led to XY male-to-female sex reversal which was evident at the cellular and transcriptional levels from early embryonic stages onwards. Conversely, in XX animals, embryonic gonads developed normally with a phenotype only observed postnatally, when adult females presented with smaller ovaries and sub-fertility. Interestingly, despite a 77% decrease in mRNA levels of *Wt1* in adult testis, immunofluorescence staining suggested an abundance of WT1 protein. This suggests that for *Wt1,* mRNA levels may not be a good proxy for protein levels, as is the case in many other genes ^69^. Additionally, while a 40% and 77% decrease in *Wt1* transcript was detected in in XX and XY adult gonads, respectively, analysis at embryonic stage E13.5 did not detect a significant decrease in mRNA levels. This raises the question as to how such a prominent phenotype can arise when a *Wt1* enhancer is deleted without any apparent change to *Wt1* mRNA expression? We propose two possible explanations: First, our data suggests this enhancer is open, and likely active, very early in gonad development (*Wt1* is expressed in coelomic epithelium/intermediate mesoderm stage from E9 ^44^ where it is important for *Nr5a1* and *Sry* expression in the developing bipotential gonads). It is possible that at E13.5, we missed an essential developmental window of activity for this enhancer, thereby missing the resulting decrease in expression caused by its deletion. However, our ATAC-seq data suggests that this enhancer continues to be accessible in E13.5 Sertoli cells and pre-granulosa cells, suggesting an alternative explanation. Here, we postulate that during embryonic development, this enhancer is mainly important for *Wt1* expression in the testis. Deletion of this enhancer in early XY embryonic gonads (causing a presumed decrease in *Wt1* expression) causes sex reversal with the emergence of ovarian cells. These ovarian cells may then use an alternative ovary-specific enhancer, allowing *Wt1* transcript levels to recover to those normally observed in XX gonads. Supporting this model, PCHi-C data from E13.5 purified Sertoli and granulosa cells identified close proximity between this enhancer and the *Wt1* gene only in Sertoli cells, but not granulosa cells.

Nevertheless, this enhancer appears to play an important role in the adult ovary, as its loss leads to female sub-fertility, indicating it may have distinct functions in different tissues and developmental stages. Similarly, we have recently shown that Enh13, a distal *Sox9* enhancer that is critical during sex determination, becomes entirely dispensable for maintaining *Sox9* expression in the testis post sex determination ^70^.

### Leydig cells are present despite the major loss of Sertoli cells

Several mouse models showing XY male-to-female sex reversal have been reported ^30,71–73^. Yet in all these cases, XY mice are fully feminized externally and develop ovaries. In contrast, XY mice lacking the *Wt1* upstream enhancer identified here are also feminized externally, have gonads with ovarian morphology, but they often retain both Müllerian-derived and Wolffian-derived reproductive tracts, including oviduct and uterus, as well as epididymis and vas deferens. Our transcriptomic and immunostaining analysis suggest that while sex reversal has occurred at the level of the supporting cells in these ovarian-like gonads, functional Leydig cells persist and are likely producing androgens. Müllerian-derived tissues regress in the presence of AMH ^56^, but this is markedly decreased in the *Wt1* enhancer-deleted XY sex reversed gonads, which explains their persistence. The more puzzling question is how Leydig cells are present in the gonads that apparently lack Sertoli cells, at least at E13.5. Normally, fetal Leydig cells are derived from *Wnt5a^+^* early steroidogenic progenitors in response to paracrine factors such as DHH and PDGF secreted by Sertoli cells between E12.5 to E15.5 ^74–76^. The expression of both genes was significantly reduced in our transcriptional profiling. Interestingly, conditional KO (cKO) of *Wt1* using a *Wt1*-Cre driver results in XY mice with a feminized external phenotype and internal reproductive tracts containing both Müllerian and Wolffian derivatives, similar to the phenotype we observed. The gonads were dysgenic, failed to express either *Sox9* or *Foxl2* ^77^ and at E14.5 XY the gonads were reduced in size, yet expressed steroidogenic genes including *Star*, *Cyp11a1* and *Hsd3b1* ^77^. Indeed, studies have suggested that WT1 may be involved in the lineage specification and balance between Sertoli/granulosa cells and steroidogenic cells ^78,79^. They showed that in absence of WT1, *Nr5a1* expression is upregulated, and somatic cells differentiate into steroidogenic cells instead of supporting cells ^78,79^. Of note, our patient No. 1 in our Supp Fig 8, which is a 46,XY DSD individual presented with both Müllerian- and Wolffian duct-derived structures.

### Tissue-specific multi-modal filtration pipeline for non-coding variant filtration in DSD

With the decrease in the cost of sequencing, a number of unresolved genetic cases are now being referred for WGS as part of research or their clinical care ^4,7^. This, however, reveals a large number of rare variants in the non-coding region, posing a significant challenge for variant curation and prioritisation. Here, we focused on small indels, and single nucleotide variants identified in WGS data of from a cohort of undiagnosed individuals with DSD. By integrating highly relevant conserved spatiotemporal chromatin accessibility datasets, together with allele frequency, as well as known sex determining genes’ TADS and quality filtering, we refined variants from 4.4 million per patient to just 92 variants on average. As a proof of concept, this allowed the identification of candidate variants in a *WT1* enhancer, as well as a homozygous SNV in a *NR5A1* enhancer that reduces enhancer activity *in vitro*. These demonstrate the feasibility of this approach, and further validation of additional candidates is currently underway.

Recently, AlphaGenome from Google DeepMind has provided some improvements on functional predictions of non-coding variants through the integration of genomic, epigenomic and transcriptomic data ^10^. In contrast, our filtration pipeline provides a tissue-specific pipeline relying on genomic data isolated from the relevant cell types, at the relevant developmental time, which we believe is more suitable for congenital conditions that affect a single tissue. Future integration of copy number variant calling from WGS or even long read sequencing promises to reveal even more candidate variants affecting dosage of enhancers, something known to cause DSD ^80^.

### The necessity and limitation of functional genomics

In this study, we identified and characterised two conserved regulatory elements associated with known gonadal genes as proof of concept: one for *Wt1/WT1,* and another for *Nr5a1/NR5A1*. In the latter, we identified a single nucleotide variant (homozygous in affected individuals) that causes a 46% reduction in enhancer activity when in front of a minimal promoter in *in vitro* assays. If this is recapitulated *in vivo*, it would suggest that this G>A change could cause a loss of almost half of *NR5A1* expression in homozygous individual. Indeed, loss of one functional *NR5A1* allele (e.g. half of the functional protein in a heterozygous null or damaging variant) is well established to cause DSD including 46,XY gonadal dysgenesis, with a number of heterozygous hypomorphic *NR5A1* variants also described. Consistently, recent work from our groups has shown that very small changes in enhancer elements, including single nucleotide variants, disrupt sex determination ^28,35^. Future work is now required to understand the mechanisms underlying gonadal dysgenesis observed in these patients carrying this single nucleotide change, evidence which would contribute to a genetic diagnosis for the family. Indeed, newly developed stem-cell derived models of the gonadal somatic cells could be used to model these variants in the future ^81–83^.

## Conclusion

Creation of a conserved regulatory atlas from mouse and human gonadal foetal somatic cells has revealed a number of putative regulatory regions associated with known gonadal genes. One such enhancer for the *Wt1* gene was found to harbour rare variants in eight individuals with DSD, and was critical for proper gonadal development in mice, providing proof-of-concept of this approach. Integration of this atlas into a multi-modal filtration pipeline has further allowed identification of candidate variants from WGS data of unexplained DSD cases including a homozygous variant upstream to the *NR5A1* gene, likely to underlie 46, XY DSD in a familial case. We propose that this tissue-specific filtration pipeline based on conserved regulatory data from the central cell types, at the appropriate time of embryonic development, may provide a means by which to identify causative variants for other congenital conditions.

## Data Availability

All data produced in the present study are available upon reasonable request to the authors

## Acknowledgements

We are very grateful to all individuals and families for participating in the research. We are grateful for Dr. Roser Vento-Tormo and Dr. Valentina Lorenzoni for providing us with the human scATAC-seq data to label the data we extracted of pre-Sertoli and pre-granulosa as well as Sertoli and granulosa cells. We thank the BIU animal facility and technicians for the help with animal maintenance. We are grateful to the BIU transgenic facility for production of CRISPR genome-edited mice. We are thankful to the Life Sciences Microscopy unit at BIU for help with imaging.

## Funding

This work is co-funded by the European Union (ERC, *EnhanceSex*, 101039928) and the Israel Science Foundation (ISF_710_2020). Views and opinions expressed are however those of the authors only and do not necessarily reflect those of the European Union or the European Research Council. Neither the European Union nor the granting authority can be held responsible for them. This research was supported by an Ideas Grant (GNT2012166) and Investigator Grant (AHS, GNT2025619) from the National Health and Medical Research Council of Australia. KLA is supported by Cybec Foundation and a Rebecca L. Cooper Fellowship. GA was supported by the Melbourne Children’s Postgraduate Health Research Scholarship. AE is supported by an Azrieli PhD Fellowship. LGAF is supported by a Lalor Fellowship. EJT was supported by a Norman Beischer Scientific Fellowship. This work is funded in part by a research grant from the European Society of Pediatric Endocrinology (to AB) and by the Agence Nationale de la Recherche (ANR; ANR-10-LABX-73 REVIVE, ANR-20-CE14-0007, ANR-23-CE14-0061 to KM and ANR-19-CE14-0022, ANR-19-CE14-0012, ANR-23-CE14-0068 to AB).

## Author contribution

NG, KLA, AHS and KM conceptualised the study. AE, IB, MR and NG conceived the mouse experiments; AE, MR and EA performed the *Wt1* enhancer experiments and analysed the data. IB and RW developed the multi-modal DSD filtration pipeline. IS analysed the RNA-seq and the human scATAC-seq data. OF, ML, YO and KB contributed to the computational filtration pipeline. Sanger sequencing was carried out by JvdB, GR and LS. Patient clinical details and genomic data were compiled and analysed by GA, JV, TT, AB, ME, ET, CC, SV, MDdS and KM. TFBS analysis and luciferase assays were carried out by GA, JvdB, BH with advice from KLA.

The manuscript was written by NG, KLA, AHS, AB and KM. All authors read and accepted the data being presented in the manuscript.

## Competing declaration

The authors declare that they have no competing interests.

## Supplementary Materials

Materials and Methods

Figs. S1 to S10

Tables S1-S6

Supp data 1-7

## Notes

### Competing Interest Statement

The authors have declared no competing interest.

### Author Declarations

Royal Childrens Hospital Melbourne Australia Ethics Committee gave ethical approval for this work

