## Supplementary files for "A conserved regulatory atlas reveals enhancers critical for mouse and human gonadal development"

**The PDF file includes:**

Materials and Methods

Figs. S1 to S10

Tables S1-S6

### Materials and Methods

#### Animal Ethics Statement

All animals were maintained with appropriate husbandry according to Bar-Ilan University ethics protocols 2305-111-1 and 2306-117-1. All mice strains were maintained on a C57BL/6J genetic background. Primers used for genotyping are listed in Table S1.

#### Design and preparation of sgRNAs mRNA

Single-guide RNAs (sgRNAs) targeting the *Wt1* enhancer region were designed using IDT CRISPR/Cas9 design tool (https://eu.idtdna.com). Each sgRNA was designed under the assumption that the Cas9 will cut 3-4 bp upstream of the PAM site.
For creating crRNA and tracrRNA duplex, crRNA and tracrRNA (IDT, cat. 224893246) were resuspended into final concertation of 200µM using Nuclease Free Duplex Buffer. crRNA and tracrRNA were mixed in 1:1 ratio, to a final complex concentration of 100µM (The two sgRNA sequences are listed in Table S2). To anneal the complex, the reaction was incubated at 95°C for 5 minutes, cooled to room temperature, and stored at -20°C for up to 2 weeks or until used.
CRISPR/Cas9 ribonucleoproteins (RNP) assembly was performed just before electroporation. RNP Mix was prepared with Opti-MEM (Thermo, cat. 31985-062), 1.2µM Cas9 Nuclease V3 500µg (IDT, cat. 1081059) and 3µM of each sgRNA. RNP Mix was incubated at room temperature for 10 minutes and then placed on ice until used.

#### Zygote harvesting and electroporation

For zygote harvesting, 4-7 weeks old C57BL6/J donor female mice were super-ovulated by administration of 5-7.5 IU of PMSG (Pregnant Mare Serum Gonadotropin; ProSpec, cat. hor-272) using intraperitoneal (i.p.) injection. Following PMSG injection (~48-50 hours), females were injected with 5-7.5 IU of hCG (Human chorionic gonadotropin; sigma cat. CG10). Subsequently, Super ovulated females were mated with C57BL6/J adult males in 1:1 ratio and checked for the presence of vaginal plug (VP) the morning after mating. Female mice displaying VP were sacrificed by cervical dislocation for oviduct dissection and zygote isolation.
The oviducts were dissected, and the ampulla nicked to release zygotes associated with surrounding cumulus cells into a homemade FHM medium containing 300 µg/ml hyaluronidase (sigma cat. H4272). Zygotes were picked using a mouth pipette and transferred to a plate with fresh 2ml FHM to subsequently pass through several FHM washes to remove cumulus cells. Next, the zygotes were moved to a plate with homemade KSOM medium. Zygotes were kept in KSOM medium in a flat-bed $CO_2$ incubator (5.3% $CO_2$, 5% $O_2$ at 37 °C) (Miri, cat. 2070047) for 30 minutes. Zygotes with the presence of polar bodies and pronucleus were isolated and washed in FHM drops followed by three washes in Opti-MEM drops and then moved to the electrode chamber containing the RNP mix.
Electroporation was performed using NEPA21 (NepaGene) electroporator in repeated pulses to first damage the zona pellucida and then favor the intracellular entry of RNPs. A large glass plate (CUY505P5 electrode with 5 mm gap) was used with 50 µl of RNP complex and 20-150 embryos. Electroporation parameters were as such: 225V poring pulse 1ms pulse width, 50 ms pulse interval and 4 repeated pulses. We then applied 20V transfer pulse, 50 ms pulse width, 50 ms pulse interval for 5 consecutive pulses. Impedance was measured before and after embryo addition as stated by the manufacturer instructions. Following electroporation, zygotes were washed with FHM and KSOM drops and incubated in a KSOM drop overnight at $CO_2$ incubator (5.3% $CO_2$, 5% $O_2$ at 37 °C). Finally, embryos were surgically transferred into oviducts of pseudo-pregnant CD1 recipient females.

**gDNA isolation and genotyping of genetically altered mice**

Genomic DNA (gDNA) was extracted from tail tissue of embryos or ear punch tissue of adult animals. For gDNA isolation from embryo samples, the PCRBIO rapid extract lysis kit was used (PCRBIO, PB15.11-S). gDNA isolation from adult earpiece included 15 min incubation at 95°C with lysis buffer composed of 10mM NaOH, 0.1mM EDTA pH 8 followed by the addition of 40mM Tris HCl pH 5.

Founder mice carrying the *Wt1_Enh* deleted allele were initially identified by PCR to detect the deleted allele, followed by Sanger sequencing to identify the breakpoints. PCR products were purified using universal DNA purification kit (TIANGENE, TI-DP214-03) or by EPPiC Fast Kit (A&A biotechnologies, 1021-500F) according to the manufacturer's instructions. Mutation analysis on the founder mice and F1 pups was done using the online TIDE tools software (https://tide.nki.nl), DECODR (https://decodr.org/) and BLAST (https://blast.ncbi.nlm.nih.gov). Two independent stable lines were established from those founders.

Founder mice carrying the deleted allele were bred to *wild type* to allow germline transmission. Two heterozygous F1 mice were bred to get homozygous mice. Once the line was established, mice were analyzed using PCR. All the PCR reactions were performed using 2X Red PCR Master Mix polymerase (PCR-BIO, cat. PB10.23-10). PCR was also performed to determine the chromosomal sex of each mouse by PCR using DreamTaq 2× PCR Master Mix (Thermo Fisher Scientific, K1082). All primer sequences are listed in Table S1.

**Timed mating and tissue preparation and imaging**

Embryos and animals carrying desired deletion were produced by crossing heterozygotes mice. Embryos were collected after timed mating at embryonic day E13.5, where day 0.5 was determined by the presence of vaginal plug (VP).

All bright field images of gonads were taken using the Nikon Eclipse Ts2R microscope with an exposure time of 10 ms and analysed using the NIS-Elements D software.

For immunostaining, gonads from embryos and 6 weeks postnatal mice were harvested and fixed overnight in 4% paraformaldehyde (Sigma Aldrich, P6148) in phosphate-buffered saline (PBS) at 4°C, washed three times with PBST (PBS with 0.1% Triton (Sigma Aldrich, 9002-93-1)) at room temperature, incubated with 20% sucrose (Fisher BioReagents, BP220-1) overnight at 4°C and then embedded in OCT (Leica, 14020108926) and stored at -80°C until further use.

**Immunofluorescence staining**

Immunofluorescence staining was performed on 10µm-thick sagittal cryostat sections (Leica, CM3050-S). Antigen retrieval was performed for embryonic and adult samples with DAKO (Target retrieval solution, Agilent, S1699) at 65°C for 30 min. Samples were then blocked in PBST containing 10% donkey serum (Sigma Aldrich, D9663) for 1 hour and incubated with primary antibodies (diluted in PBST containing 1% donkey serum) overnight at 4ºC (All primary and secondary antibodies as well as dyes used are listed in Table S3). Following three PBST washes, secondary antibodies were added for 1 hour at room temperature (RT). Slides were then washed, dried, and mounted (Polysciences 18606-20). All immunofluorescence slides were also stained with 4',6-diamidino-2-phenylindole (DAPI; Invitrogen, D1306) to visualize nuclear DNA. Images were obtained with a Leica Microsystems SP8 confocal microscope.

**RNA isolation, cDNA preparation, and Quantitative Real-Time Polymerase Chain Reaction (qRT-PCR)**

Total RNA was extracted from 6w gonads using EURX GeneMATRIX Universal RNA Purification Kit (EURX, E3598). RNA yield was quantified using a NanoDrop spectrophotometer. For adult samples, 500 ng of RNA was taken from 6 weeks old gonads for cDNA preparation using the qScript cDNA Synthesis Kit (QuantaBio 95047-100-2) according to the manufacturer's instructions. qRT-PCR reactions were performed in duplicate using qPCR-PerfeCTa SYBR Green FastMix (95074-012-2, QuantaBio) with 140 nM each of forward and reverse primers (Table S4) and analyzed on the QuantStudio 1 Real-Time PCR System (Thermo Scientific). Analysis was done using Comparative CT ($2^{-\Delta\Delta CT}$) technique and expression is relative to the house keeping gene, *Hprt.* Statistical analyses were carried out using Prism 10 software (GraphPad) using one-way ANOVA test followed by Dunnett's post-test. P value < 0.05 was considered statistically significant. The number of gonads analyzed at each stage is depicted in the graphs.

**RNA-seq and library prep**

Gonad pairs of E13.5 mice were dissected, separated from the mesonephros and snap-frozen in liquid nitrogen. RNA was extracted using Qiagen RNeasy Plus micro kit (Qiagen, 74034). RNA yield was quantified with Qubit RNA HS (Invitrogen, Q32852). Quality of the RNA was analyzed using High sensitivity RNA ScreenTape Assay (Agilent, 5067-5579). 200 ng of RNA was used and transcribed to cDNA. Libraries were prepared using Poly(A) mRNA Magnetic Isolation Module (NEB, E7490S), along with NEBNext UltraExpress RNA library prep kit (NEB, E3330S) and cleaned using AMPure beads (Beckman coulter, Bc-a63881) according to manufacturer's instruction. Unique indexes were added to each sample using ++NEBNext Multiplex Oligos for Illumina kit (NEB-E6440S) according to manufactures instruction and cleaned using AMPure beads. cDNA and library concentrations were analysed using Qubit ds HS Assay Kit (Invitrogen, 2326054) and sample size distribution using TapeStation with high sensitivity D1000 tape (Agilent, 5067-5585). Samples were pooled together to create a 2nM library and sequenced with 60bp paired end (PE) reads on an Illumina NextSeq 2000 platform at the Kanbar core facility unit, Bar-Ilan University, generating ~40-50 million reads per sample.

**RNA-seq mapping and differential expression analysis**

FastQ files were processed using nf-core/rnaseq pipeline v3.12.0. Briefly, read quality controls were performed with FastQC. Sequencing adapters were removed with TrimGalore!. Reads were mapped on the mm10/GRCm38 reference genome from Gencode (M25) with STAR. Gene quantification as read counts and TPM (Transcript Per Million) was obtained using RSEM. Read count matrix was filtered to exclude lowly expressed genes (genes with less than 10 reads and/or TPM value less than 10). Non-protein-coding genes were filtered out in the subsequent analysis. Sample correlation (Spearman) and PCA were performed with R (corr and prcomp functions) using the filtered read counts normalized by library size (sizeFactor) with DESeq2. Differential expression analysis was performed on the filtered read count matrix using DESeq2 using "LRT" (Likelihood Ratio Test). Genes with log2(FoldChange) < -0.5 and log2(FoldChange) > 0.5, as well as an adjusted *p*-value < 0.01 were considered as differentially expressed.

Gene expression values (filtered read count matrix normalized by library size) of the differentially expressed genes were transformed as z-scores, clustered into groups according to their expression profiles using hclust and "ward.D2" method and represented as heatmap using ComplexeHeatmap. The optimal number of clusters were assessed using the best.cutree function from the Jlutils R package (https://github.com/larmarange/Jlutils), with a minimum of

four possible clusters. Samples were ordered according to the hierarchical clustering ("ward.D2") and split into three groups.
The code used to analyse the RNA-seq data is available in Github: https://github.com/IStevant/Wt1_enhancer_deletion. RNA-seq data has been deposited in the Gene Expression Omnibus under accession numbers GSE324087 (Review access token: ixqbqkcmrvyzjof).

**Fertility tests**
For fertility tests, adult females (XX *Wt1_Enh*$^{+/+}$ or XX *Wt1_Enh*$^{\Delta931/\Delta931}$) were single caged with adult *Wt1_Enh*$^{+/+}$ males for a total period of 6 months. During this time period, the number of litters born as well as number of pups born per litter were documented.

**Patients Ethics Approval**
The study was approved under HREC22097 (Royal Children's Hospital, Melbourne, Australia) as well as the French ethical committee (2014/18NICB-registration number IRB00003835). All participants signed or had a parent/guardian sign informed consent and DNA from blood and clinical data were collected as part of routine care. Participants have been deidentified.

**Whole genome sequencing (WGS) of DSD individuals**
**WGS Melbourne cohort:** All individuals were previously negative for diagnostic findings in microarray and whole exome sequencing (with DSD panel applied). WGS was performed at the Victorian Clinical Genetics Services, Melbourne, Australia with Illumina DNA PCR-free at 30x, with Illumina Novaseq 6000 sequencing. **NR5A1 variant family**: WGS was carried out for both affected children, parents and one unaffected brother. The alleles were confirmed in the second unaffected brother using Sanger sequencing.
**WGS Pasteur cohort**: All individuals were previously negative for pathogenic variants in genes known to be associated with DSD by whole exome sequencing. WGS was perform by Bejing Genomics Institute at 30x using the PCR-free library sequencing platform DNBSEQ.

**Cloning**
Both "ENCODE" and "ATAC" genomic fragments were amplified from patient genomic DNA and unaffected family members using standard Phusion polymerase protocols and cloned into the EcoRV/XhoI restriction enzyme sites of the luciferase reporter vector pGL4.10[luc2] (Promega, E6651). While the EnhP129 enhancer fragment was cloned into MCS of the pGL4.26[luc2/minP/Hygro] (Promega, E8441) using the same restriction sites as above. Fragments were cloned in the corresponding genomic orientation to drive luciferase reporter gene expression. (Details of primer and vectors, Table S5, Table S6)

**Sanger sequencing for validating WGD data**
The WGS identified variants were verified using the Sanger sequencing BigDye Terminator 3.1 from Applied Biosystems, primers listed in Table S5 and read on AB 3730xl instrument (service provided by AGRF).
Undiagnosed 46, XY individuals with gonadal dysgenesis for whom no WGS was available were screened for variants within the *WT1* enhancer region using Sanger sequencing as described above (primers listed in Table S5).

**Luciferase Assays**
HEK293T and COS7 cells were grown in DMEM (Dulbecco's Modified Eagle Medium) +10% FBS (fetal bovine serum). Cells were seeded at 70-80% confluency in a 96 well plate for 4-6 hours, and each well was transfected with a combination of reporter plasmid (75ng) with 2.5ng

of Renilla luciferase as a control, using Lipofectamine 2000 (ThermoFisher, Cat No. 11668019) for 24 hours.
For all luciferase assays, we normalised Firefly luciferase values to Renilla luciferase values (transfection efficiency control) and then plotted them relative to the empty luciferase vector, or relative to the *wild type* control for variants. All luciferase experiments were carried out in at least 3 biological replicates (independent experiments) each with 3 technical replicates (wells). We calculated standard error of the mean (SEM) and either two-tailed Students *t*-test or ANOVA using Graphpad Prism V10.

**Whole-genome variant normalisation and annotation**
Whole-genome sequencing (WGS) data were provided as per-proband (Single/trio/family) variant call files (VCF). Each VCF was aligned to the human reference genome build GRCh38/hg38, compressed and indexed before processing. To ensure that each record represented a single, left-aligned variant, the files were normalised using **bcftools 1.19-69-g466ceaeb**. The bcftools norm command left-aligned insertions and deletions, checked that the reference allele matched the reference genome, and split multi-allelic records into single-variant rows. Variants outside of the genomic regions of interest were removed using bcftools' region-filtering options; only variants falling within the merged human and mouse gonadal ATAC-seq peaks were retained.
Population allele-frequency annotation was obtained from **gnomAD v4** [1]. For each variant, the GroupMax filtering allele frequency (popmax AF) was extracted from gnomAD. Functional genomic annotations were added using the **NCBI RefSeq** gene models [2]. To identify variants located in repetitive DNA, the normalised VCFs were screened with **RepeatMasker** (http://www.repeatmasker.org).

**DSD gene regions**
A list of DSD-related genes (Supp Data 6) was used to define the regions of interest. Gene coordinates were obtained from NCBI RefSeq [2]. TAD boundaries were taken from the human TAD database [3]. For each gene, the longest transcript was selected, and its genomic range was intersected with the corresponding TAD. When no TAD was available, a ±1.5 Mb flanking region around the gene coordinates was used.

**Creating Genomic region of interest**
We defined cross-species gonadal accessible chromatin regions by integrating human and mouse ATAC-seq peaks on a common human reference genome (hg38). First, human gonadal scATAC-seq [4] peaks (hg38) were sorted and merged using bedtools v2.27.1 [5] (mm10) were similarly sorted and merged with bedtools. Third, the merged mouse intervals were lifted over from mm10 to GRCh38/hg38 using liftOver command-line utility from the UCSC Genome Browser suite [6] with the mm10ToHg38.over.chain.gz chain file, enabling multiple mappings and using a minimum match ratio of 0.1; post-conversion, multiply mapped and size-inconsistent intervals were filtered as per our utility script to retain one-to-one, size-concordant mappings. Finally, the hg38 human set and the hg38-converted mouse set were concatenated and merged with bedtools to generate a unified, non-redundant set of genomic regions of interest on hg38 for downstream analyses.

**ID format for merged human–mouse segments**
Segment identifiers follow a conditional schema: *<human>hg38<mouse>* when both species support the interval; *<human>* when only human support exists, and *hg38_<mouse>* when the interval is found only in converted mouse data.

The human part is a dot-separated block of cell-type codes in canonical order- GR (granulosa), PG (pre-granulosa), PS (pre-sertoli), SE (Sertoli) with the terminal code suffixed by a serial (e.g., SE40369).
The mouse part encodes the mm10 segments converted to hg38 contribution as *XY<stages>[XX<stages>]. <index>*, where *<stages>* are embryonic stage numbers compacted by removing the ".5".
Thus, GR.PG.PS.SE40369_hg38_XY11.12.13.15XX11.13.15.55589 matches both-species form; GR.PG.SE17 illustrates human-only; and hg38_XY11.12.13XX11.13.15.12345 illustrates mouse-only.

**Variant filtering from WGS data**

The normalised VCF files, DSD gene regions, merged human–mouse gonadal ATAC-seq regions, RepeatMasker annotations, and sample metadata were uploaded into a PostgreSQL v18 database. A structured SQL query was used to generate a quality-filtered variant table by retaining only variants overlapping the predefined DSD and gonadal accessible chromatin regions and excluding variants located within repetitive regions annotated by RepeatMasker. Quality-control filtering was then applied, requiring variants present in gnomAD v4 to pass gnomAD quality filters, while all variants were additionally required to meet internal sequencing quality thresholds of $10 < DP < 100$ and $GQ > 80$. Cohort-level filtering was performed using one representative sample per family, excluding variants with GQ or DP values below the cohort median and variants present in more than 30% of unrelated individuals. The resulting table included cohort-level summary variables for each variant, including the total number of carriers, the number of proband carriers, and the number of non-proband carriers.

**Genomic constraint scores**

Genomic constraint scores (Z scores) were extracted from the computed values published in Supplementary Data 3 file from Chen et al. [1]. Z scores for the gene regions of interest: *WT1*enhancer (chr11:32499150-32500999) and *NR5A1* enhancer (chr9:124507636-124507946) were extracted using Python v3.14 for overlapping 1 kb sliding-windows with 100 bp steps. The weighted average Z score was calculated for regions overlapping the enhancer coordinates.

**Data and materials availability**

All data is available in the manuscript or the supplementary materials. RNA-seq data have been deposited in the Gene Expression Omnibus under accession numbers GSE324087 (Review access token: ixqbqkcmrvyzjof). The code produced to analyze the RNA-seq data is available on GitHub: https://github.com/IStevant/Wt1_enhancer_deletion. The code produced to analyse and integrate that ATAC-seq datasets along with the multi-model filtration pipeline is available on GitHub: https://github.com/Toozig/bed_merger

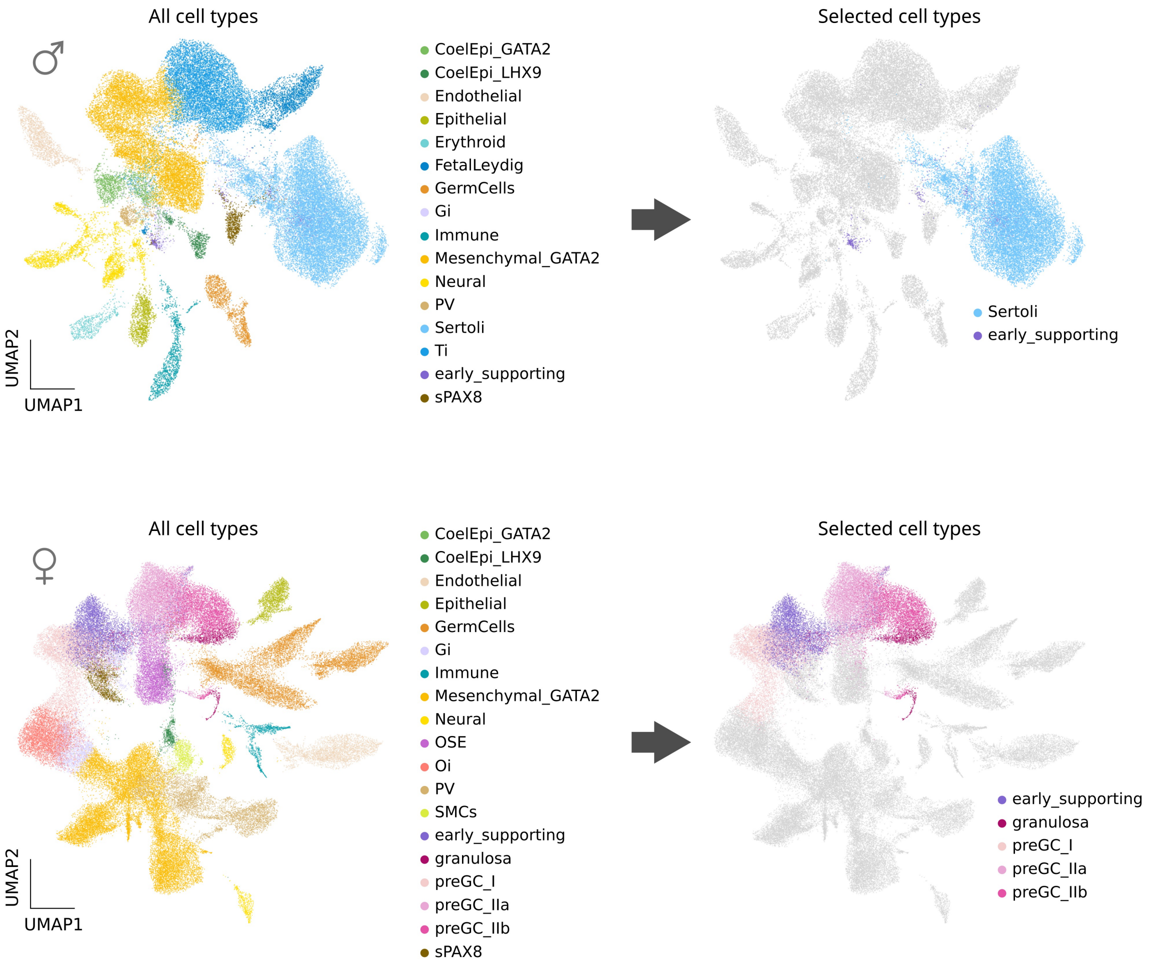

**Supplementary Figure 1. Putative regulatory elements in human embryonic gonadal supporting cell precursors as well as Sertoli and granulosa cells, extracted from scATAC-seq.** Top left shows the original UMAP of cell lineages in the human male scATAC-seq (n = 52,285). Bottom left shows UMAP of cell lineages in the human female scATAC-seq (n = 84,631). Original data relies on Garcia-Alonso et al., *Nature* 2022. Right panels show the cell populations from which scATAC-seq peaks were extracted for downstream analyses (see Supp data 1) in males (top) and females (bottom) embryonic human gonads. CoelEpi, coelomic epithelium; Endo, endothelial; Epi, epithelial; F. Leydig, fetal Leydig; Gi, gonadal interstitial; Mesen, mesenchymal; Oi, ovarian interstitial; OSE, ovarian surface epithelium; preGC, pregranulosa cells; PV, perivascular; sPAX8, supporting PAX8+; Ti, testicular interstitial; SMC, smooth muscle cell.

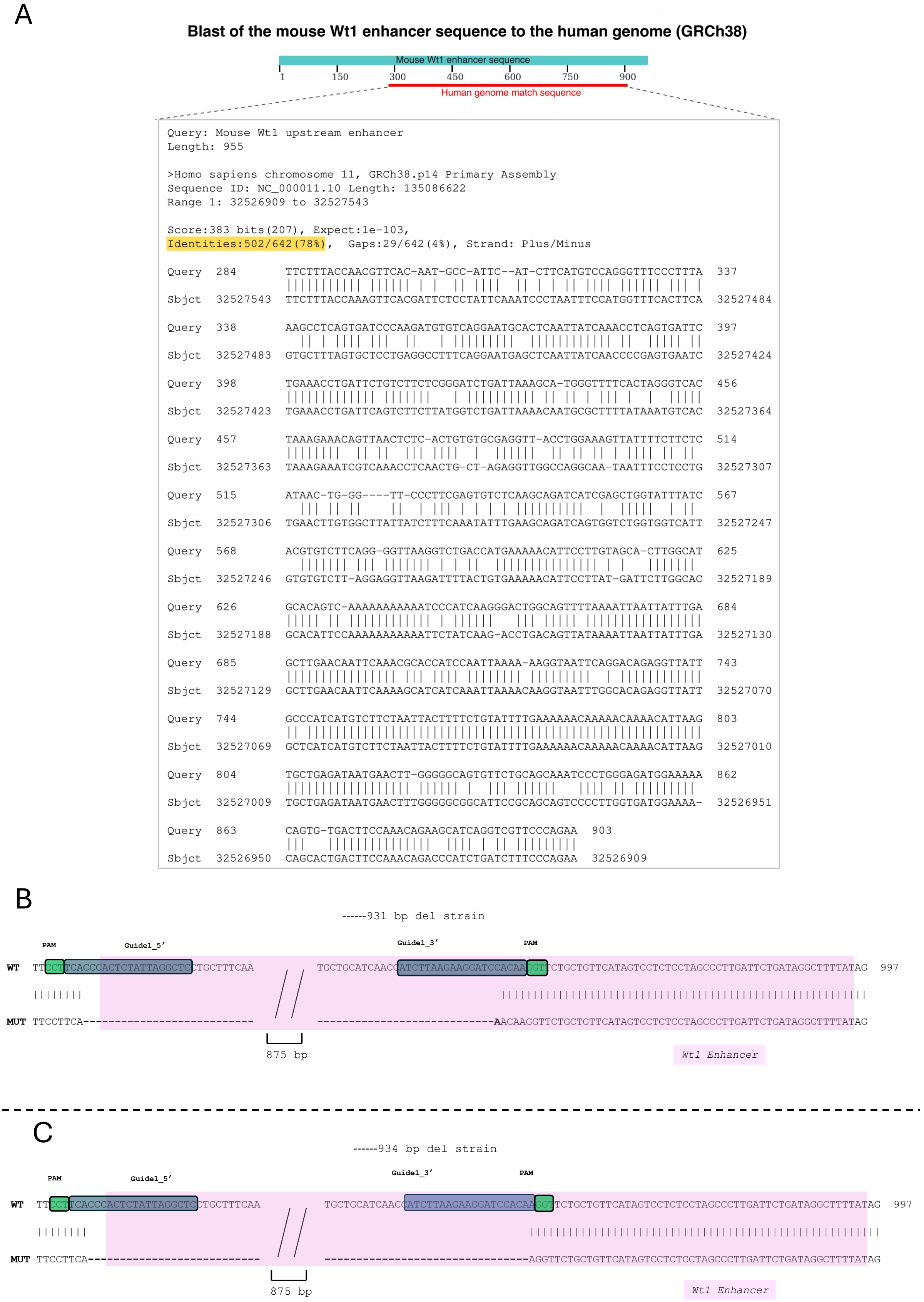
A
Blast of the mouse Wt1 enhancer sequence to the human genome (GRCh38)
Mouse Wt1 enhancer sequence
1 150 300 450 600 750 900
Human genome match sequence
Query: Mouse Wt1 upstream enhancer
Length: 955
>Homo sapiens chromosome 11, GRCh38.p14 Primary Assembly
Sequence ID: NC_000011.10 Length: 135086622
Range 1: 32526909 to 32527543
Score:383 bits(207), Expect:1e-103,
Identities:502/642(78%), Gaps:29/642(4%), Strand: Plus/Minus
Query 284 TTCTTTACCAACGTTCAC-AAT-GCC-ATTC--AT-CTTCATGTCCAGGGTTTCCCTTTA 337
Sbjct 32527543 TTCTTTACCAAAGTTCACGATTCTCCTATTCAAATCCCTAATTTCCATGGTTTCACTTCA 32527484
Query 338 AAGCCTCAGTGATCCCAAGATGTGTCAGGAATGCACTCAATTATCAAACCTCAGTGATTC 397
Sbjct 32527483 GTGCTTTAGTGCTCCTGAGGCCTTTCAGGAATGAGCTCAATTATCAACCCCGAGTGAATC 32527424
Query 398 TGAAACCTGATTCTGTCTTCTCGGGATCTGATTAAAGCA-TGGGTTTTCACTAGGGTCAC 456
Sbjct 32527423 TGAAACCTGATTCAGTCTTCTTATGGTCTGATTAAAACAATGCGCTTTTATAAATGTCAC 32527364
Query 457 TAAAGAAACAGTTAACTCTC-ACTGTGTGCGAGGTT-ACCTGGAAAGTTATTTTCTTCTC 514
Sbjct 32527363 TAAAGAAATCGTCAAACCTCAACTG-CT-AGAGGTTGGCCAGGCAA-TAATTTCCTCCTG 32527307
Query 515 ATAAC-TG-GG----TT-CCCTTCGAGTGTCTCAAGCAGATCATCGAGCTGGTATTTATC 567
Sbjct 32527306 TGAACTTGTGGCTTATTATCTTTCAAATATTTGAAGCAGATCAGTGGTCTGGTGGTCATT 32527247
Query 568 ACGTGTCTTCAGG-GGTTAAGGTCTGACCATGAAAAACATTCCTTGTAGCA-CTTGGCAT 625
Sbjct 32527246 GTGTGTCTT-AGGAGGTTAAGATTTTACTGTGAAAAACATTCCTTAT-GATTCTTGGCAC 32527189
Query 626 GCACAGTC-AAAAAAAAAAATCCCATCAAGGGACTGGCAGTTTTAAAATTAATTATTTGA 684
Sbjct 32527188 GCACATTCCAAAAAAAAAAAATTCTATCAAG-ACCTGACAGTTATAAAATTAATTATTTGA 32527130
Query 685 GCTTGAACAATTCAAACGCACCATCCAATTAAAA-AAGGTAATTCAGGACAGAGGTTATT 743
Sbjct 32527129 GCTTGAACAATTCAAAAGCATCATCAAATTAAAACAAGGTAATTTGGCACAGAGGTTATT 32527070
Query 744 GCCCATCATGTCTTCTAATTACTTTTCTGTATTTTGAAAAAACAAAAACAAAACATTAAG 803
Sbjct 32527069 GCTCATCATGTCTTCTAATTACTTTTCTGTATTTTGAAAAAACAAAAACAAAACATTAAG 32527010
Query 804 TGCTGAGATAATGAACTT-GGGGGCAGTGTTCTGCAGCAAATCCCTGGGAGATGGAAAAA 862
Sbjct 32527009 TGCTGAGATAATGAACTTTGGGGGCGGCATTCCGCAGCAGTCCCCTTGGTGATGGAAAA- 32526951
Query 863 CAGTG-TGACTTCCAAACAGAAGCATCAGGTCGTTCCCAGAA 903
Sbjct 32526950 CAGCACTGACTTCCAAACAGACCCATCTGATCTTTCCCAGAA 32526909
B
------931 bp del strain
PAM
Guide1_5'
Guide1_3'
PAM
WT TTCCTTCACCCACTCTATTAGGCTCCTGCTTTCAA
TGCTGCATCAACCATCTTAAGAAGGATCCACAAGGTCCTGCTGTTCATAGTCCTCTCCTAGCCCTTGATTCTGATAGGCTTTTATAG 997
MUT TTCCTTCA---------------------------
----------------------------AACAAGGTTCTGCTGTTCATAGTCCTCTCCTAGCCCTTGATTCTGATAGGCTTTTATAG
875 bp
Wt1 Enhancer
C
------934 bp del strain
PAM
Guide1_5'
Guide1_3'
PAM
WT TTCCTTCACCCACTCTATTAGGCTCCTGCTTTCAA
TGCTGCATCAACCATCTTAAGAAGGATCCACAAGGTTCTGCTGTTCATAGTCCTCTCCTAGCCCTTGATTCTGATAGGCTTTTATAG 997
MUT TTCCTTCA---------------------------
--------------------------------AGGTTCTGCTGTTCATAGTCCTCTCCTAGCCCTTGATTCTGATAGGCTTTTATAG
875 bp
Wt1 Enhancer

**Supplementary Figure 2. Conservation of the *Wt1* upstream enhancer and mouse strains containing CRISPR/Cas9-mediated deletion of the *Wt1* enhancer.** (A) Blast sequence comparison between the mouse and human Wt1 upstream enhancer. (B-C) Sequence alignment of *Wt1_Enh wild type* and *Wt1_Enh deleted* alleles from two independently generated *Wt1_Enh* deletion lines: a 931 bp deletion +A insertion (B) and a 934 bp deletion (C). 5' and 3' guide RNA (gRNA) used to target the sequences are highlighted in blue, and adjacent PAM sequences are shown in green. The pink shaded region indicates the *Wt1_Enh* interval. Dashed lines in the mutant alleles represent the excised genomic sequence. Double slashes (//) denote part of the deleted segment for clarity.

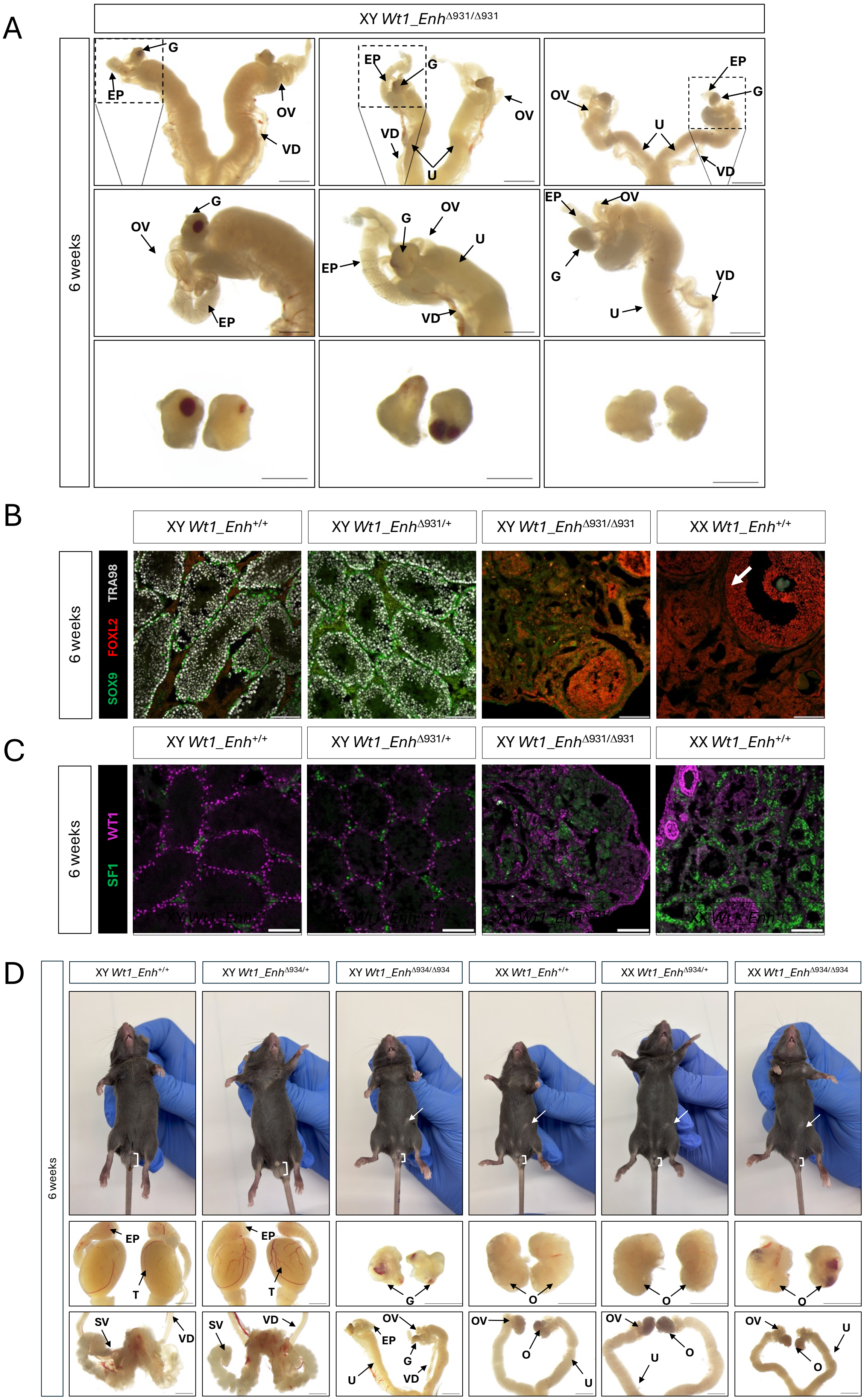
A
XY Wt1_Enh$^{\Delta931/\Delta931}$
6 weeks
G
EP
OV
VD
U
B
XY Wt1_Enh$^{+/+}$
XY Wt1_Enh$^{\Delta931/+}$
XY Wt1_Enh$^{\Delta931/\Delta931}$
XX Wt1_Enh$^{+/+}$
SOX9 FOXL2 TRA98
C
WT1
SF1
D
XY Wt1_Enh$^{\Delta934/+}$
XY Wt1_Enh$^{\Delta934/\Delta934}$
XX Wt1_Enh$^{\Delta934/+}$
XX Wt1_Enh$^{\Delta934/\Delta934}$
T
O
SV

1

**Supplementary Figure 3. Extended characterization of the feminized XY *Wt1_Enh-/-* mice.** (A) Magnified brightfield images of the variable phenotype of XY *Wt1_Enh*$^{Δ931/Δ931}$ mice. Scale bars represent 1000 µm. (B-C) Immunostaining of 6-week-old XY *Wt1_Enh*$^{+/+}$, *Wt1_Enh*$^{Δ931/+}$, *Wt1_Enh*$^{Δ931/Δ931}$ and XX *Wt1_Enh*$^{+/+}$ mice. Gonads were stained for (B) Sertoli-marker SOX9 (green), granulosa-marker FOXL2 (red) and germ cell marker TRA98 (grey), and (C) for bipotential somatic cell markers WT1 (magenta) and SF1 (green). Scale bars represent 100 µm. (D) Brightfield images of 6 weeks-old external and internal genitalia of an independent mouse line carrying 934 bp deletion, eliminating most of the *Wt1* enhancer. Presented are XY and XX *Wt1_Enh*$^{+/+}$, *Wt1_Enh*$^{Δ934/+}$, *Wt1_Enh*$^{Δ934/Δ934}$ mice. Brackets indicate anogenital (AG) distance; white arrows indicate nipples. Scale bars represent 2000 µm (testis and reproductive tract) and 1000 µm (gonads). Abbreviations (A,D) represent structures of the reproductive system: T, testis; VD, vas deferens; SV, seminal vesicles; EP, epididymis; T, testis; G, gonad; OV, oviduct; O, ovary; U, uterus.

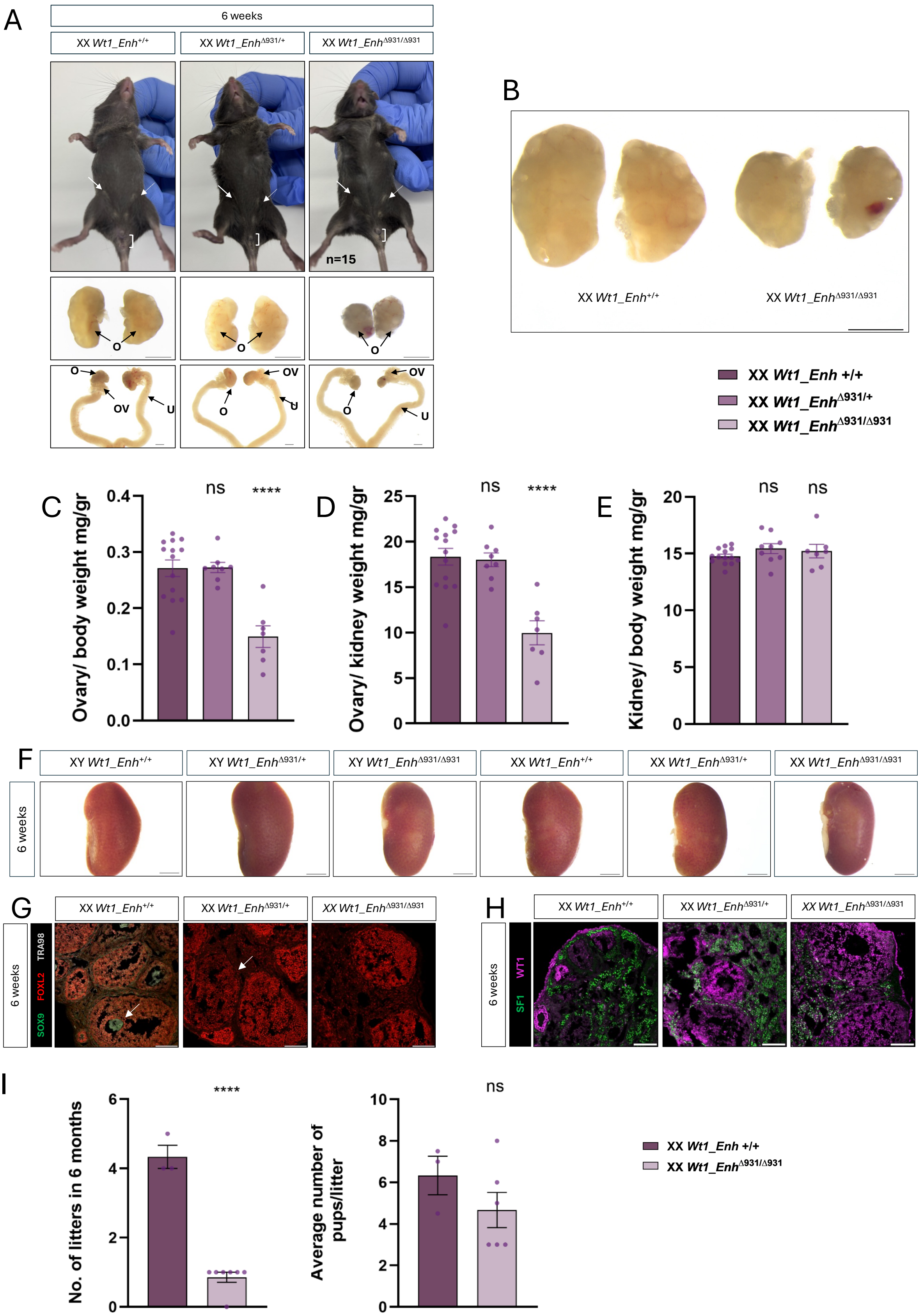

**Supplementary Figure 4. Adult XX *Wt1_Enh*$^{\Delta931/\Delta931}$ mice display reduced ovarian size and major sub-fertility.** (A) Brightfield images of external genitalia and internal reproductive tracts of 6-weeks-old XX *Wt1_Enh*$^{+/+}$, *Wt1_Enh*$^{\Delta931/+}$, *Wt1_Enh*$^{\Delta931/\Delta931}$ mice. Brackets indicate anogenital (AG) distance; white arrows indicate nipples. Scale bars represent 2000 µm (reproductive tract) and 1000 µm (ovaries). Abbreviations: O, ovary; OV, oviduct; U, uterus. (B) Representative brightfield images of XX *Wt1_Enh*$^{+/+}$ and *Wt1_Enh*$^{\Delta931/\Delta931}$ ovaries shown side by side for size comparison. Scale Bar represents 1000 µm. (C-E) Quantification of ovary weight in 6-weeks-old XX *Wt1_Enh*$^{+/+}$, *Wt1_Enh*$^{\Delta931/+}$, *Wt1_Enh*$^{\Delta931/\Delta931}$ mice. Ovary weight was normalized to (C) body weight or (D) kidney weight. (E) Kidney weight normalized to body weight, showing no significant differences among genotypes. Each dot represents the average weight of both ovaries, or kidneys, of an individual mouse; Data are presented as mean ± SEM. Statistical analysis was performed using one-way ANOVA followed by Dunnett's post hoc test in Prism 10 software where samples were compares to *Wt1_Enh*$^{+/+}$. **P < 0.05, **P < 0.01, ***P < 0.001, and ****P < 0.0001,* ns- not significant. P values below 0.05 were considered statistically significant. (F) Bright field images of 6-weeks-old kidneys of XY and XX *Wt1_Enh*$^{+/+}$, *Wt1_Enh*$^{\Delta931/+}$, *Wt1_Enh*$^{\Delta931/\Delta931}$ mice. Scale Bar represents 2000 µm. (G-H) Immunostaining of 6-weeks-old XX gonads. XX *Wt1_Enh*$^{+/+}$, *Wt1_Enh*$^{\Delta931/+}$, *Wt1_Enh*$^{\Delta931/\Delta931}$ gonads were stained for (G) the Sertoli-marker SOX9 (green), granulosa-marker FOXL2 (red) and germ cells marker TRA98 (grey) and for (H) bipotential somatic cell markers WT1 (magenta) and SF1 (green). Scale bars represent 100 µm. (I) Fertility tests of XX *Wt1_Enh*$^{\Delta931/\Delta931}$ mice compared with *Wt1Enh*$^{+/+}$ controls. Each dot represents an individual female. The left graph shows the average number of litters per born per female over a period of 6 months, and the right graph shows the number of pups born per litter. Data are presented as mean ± SEM. Statistical analysis was performed using unpaired Mann–Whitney test in GraphPad Prism 10. **P < 0.05, **P < 0.01, ***P < 0.001, and ****P < 0.0001,* ns- not significant. *P* values below 0.05 were considered statistically significant.

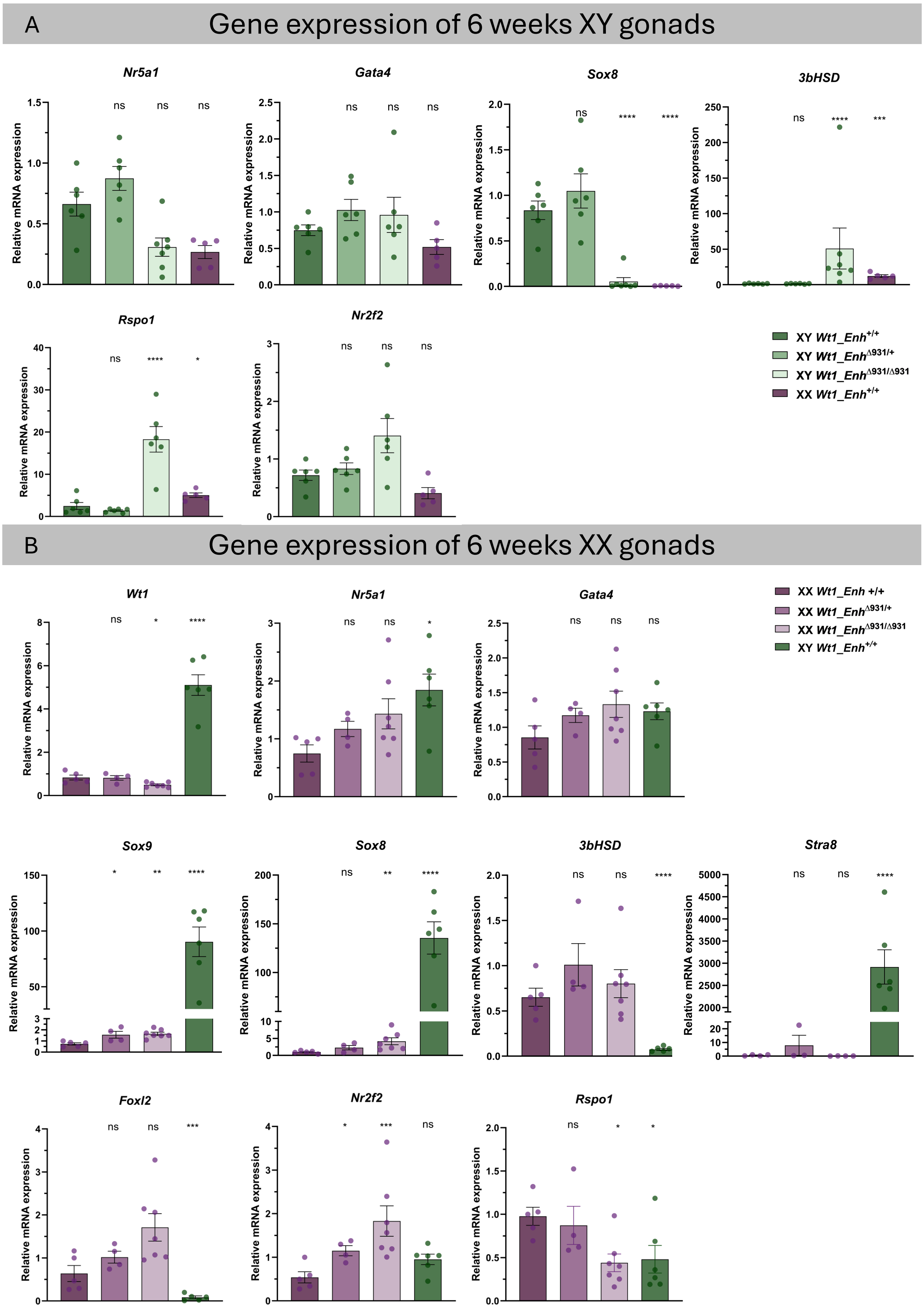

**Supplementary Figure 5. Gene expression analysis of 6 weeks-old gonads of *Wt1_Enh*$^{\Delta931/\Delta931}$ mice.** Real-time quantitative PCR analysis of genes involved in male and in female gonadal development in 6-week-old gonads of XY mice (A) and XX mice (B). Data are presented as mean $2^{-\Delta\Delta Ct}$ ± SEM and were normalized to the housekeeping gene *Hprt*. Sample size is indicated as dots representing the number of individual mice. Statistical significance was determined using one-way ANOVA followed by Dunnett's post hoc test. **P < 0.05, **P < 0.01, ***P < 0.001,* and *****P < 0.0001,* ns- not significant. P values below 0.05 were considered statistically significant. In A, all samples are compared to the XY *Wt1_Enh+/+*. In B, all samples are compared to the XX *Wt1_Enh*$^{+/+}$.

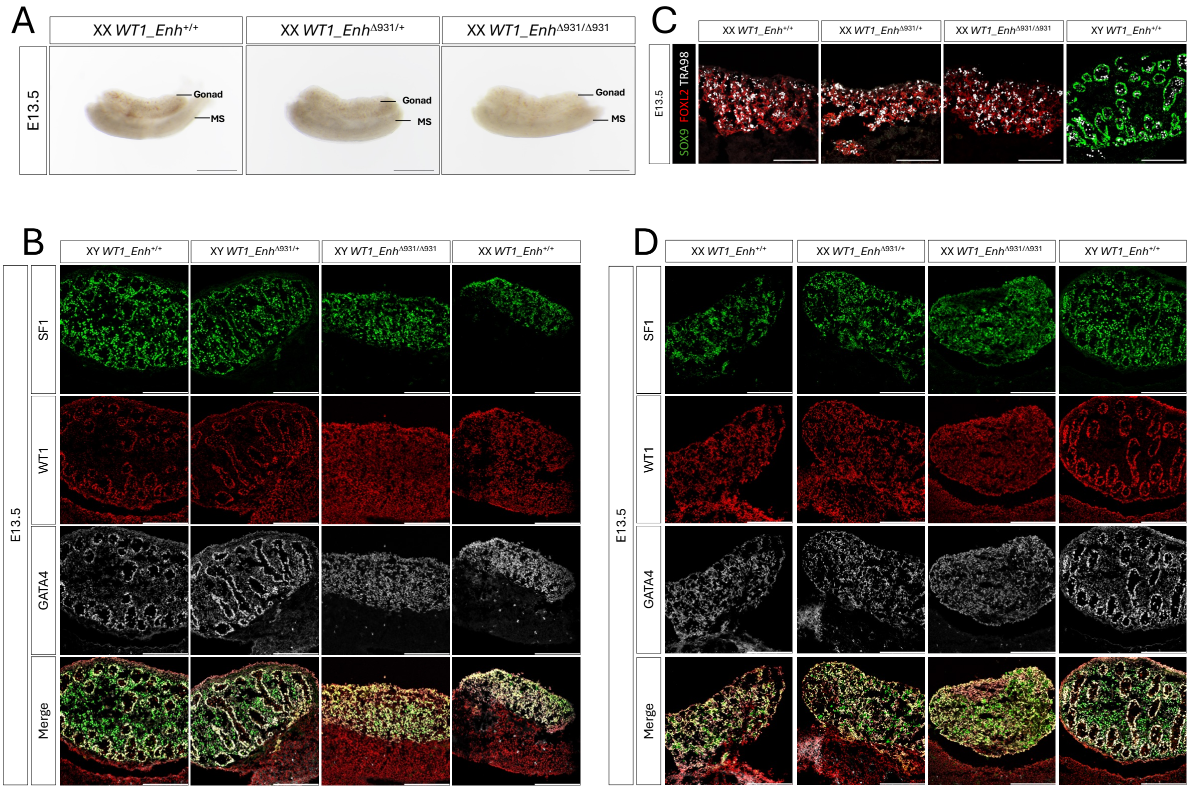

**Supplementary Figure 6. Extended analysis of embryonic gonadal phenotypes in *Wt1_Enh*$^{\Delta931/\Delta931}$ mice**. (A) Bright field images of gonads from E13.5 XX *Wt1_Enh*$^{+/+}$, *Wt1_Enh*$^{\Delta931/+}$, *Wt1_Enh*$^{\Delta931/\Delta931}$ embryos. (B,D) Immunostaining of E13.5 XY (B) and XX (D) *Wt1_Enh*$^{+/+}$, *Wt1_Enh*$^{\Delta931/+}$, *Wt1_Enh*$^{\Delta931/\Delta931}$ gonads. Gonads were stained for bipotential somatic cell markers SF1 (green), WT1 (red) and GATA4 (grey). (C) Immunostaining of E13.5 XX *Wt1_Enh*$^{+/+}$, *Wt1_Enh*$^{\Delta931/+}$, *Wt1_Enh*$^{\Delta931/\Delta931}$ and XY *Wt1_Enh*$^{+/+}$ embryos. Gonads were stained for Sertoli-marker SOX9 (green), granulosa-marker FOXL2 (red) and germ cell marker TRA98 (grey). Scale bars represent 500 µm in (A) and 200 µm in (B–D).

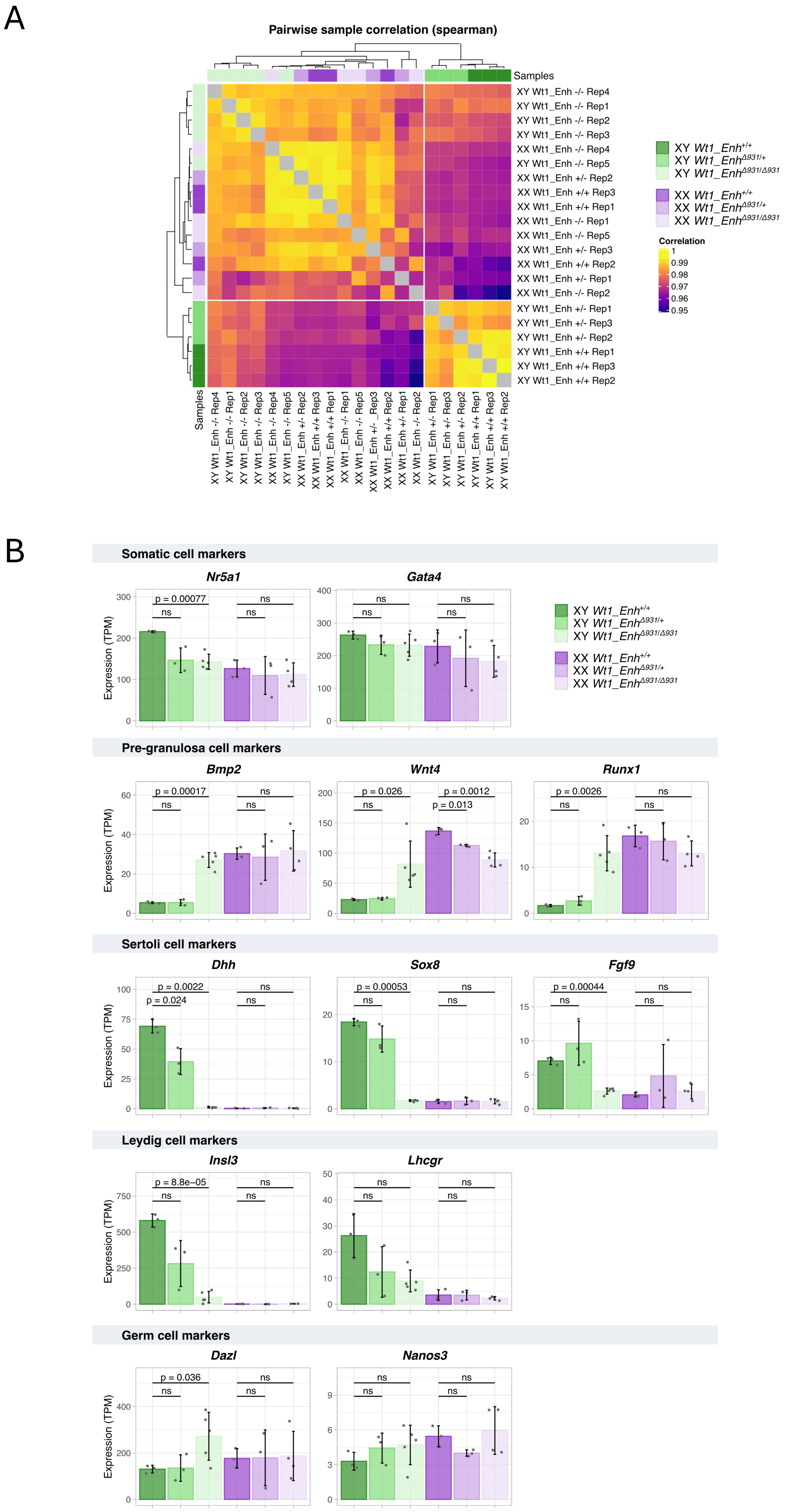
A
Pairwise sample correlation (spearman)
Samples
Correlation
1
0.99
0.98
0.97
0.96
0.95
B
Somatic cell markers
Nr5a1
Gata4
Pre-granulosa cell markers
Bmp2
Wnt4
Runx1
Sertoli cell markers
Dhh
Sox8
Fgf9
Leydig cell markers
Insl3
Lhcgr
Germ cell markers
Dazl
Nanos3
Expression (TPM)

**Supplementary Figure 7. Transcriptomic analysis of E13.5 embryonic gonads of *Wt1_Enh*$^{\Delta 931/\Delta 931}$ embryos.** (A) Pearson correlation of the transcriptomes from XY and XX *Wt1_Enh*$^{+/+}$, *Wt1_Enh*$^{\Delta 931/+}$ and *Wt1_Enh*$^{\Delta 931/\Delta 931}$ E13.5 gonads. Samples are grouped by similarity using unsupervised hierarchical clustering as shown by the dendrogram on the top and the left-hand side of the heatmap. (B) Expression profiles of well-known gonadal marker genes including markers of somatic cells, Sertoli cells, pre-granulosa cells, Leydig cells and germ cells. Expression values are expressed as TPM (Transcript per million), the bars represent the median of expression, each point represent a single biological replicate. Error bars represent the standard deviation across the replicates. Statistical significance was determined using one-way ANOVA followed by Dunnett's post hoc test.

A

| ID | Chr | Position | Ref | Alt | rsID | AFpopmax | TAD | Distance from nearest DSD gene | Inheritance | Phenotype | Ancestry |
|---|---|---|---|---|---|---|---|---|---|---|---|
| 1 | 11 | 32,499,247 | T | TG | rs123827277 | 0.0000147 | *WT1* | 63708 | maternal | XY DSD | - |
| 2 | 11 | 32,499,312 | A | G | rs192741756 | 0.0005611 | *WT1* | 63775 | (*)- | XY T/OTDSD | Brazil |
| 3 | 11 | 32,499,568 | G | C | novel | 0.0 | *WT1* | 99168 | maternal | XX DSD | North Africa |
| 4 | 11 | 32,499,739 | T | C | novel | 0.0 | *WT1* | 64200 | - | XX DSD | North Africa |
| 5 | 11 | 32,500,052 | C | T | novel | 0.0 | *WT1* | 99652 | - | XY GD | South East Asia |
| 6 | 11 | 32,500,193 | G | A | rs1854184058 | 0.00002413 | *WT1* | 101588 | - | XY GD | European |
| 7 | 11 | 32,500,228 | T | A | rs1242545063 | 0.00006547 | *WT1* | 102072 | maternal | XY GD | European |
| 8 | 11 | 32,500,249 | G | A | rs1854184441 | 0.00004880 | *WT1* | 101104 | - | XY GD | European |

* Variant segregates with the affected sibling and is absent in the unaffected mother. The pedigree suggests paternal transmission, as an affected uncle has been identified (DOI: 10.1007/BF02281867); however, the father's sample was not available to confirm paternal inheritance.

B

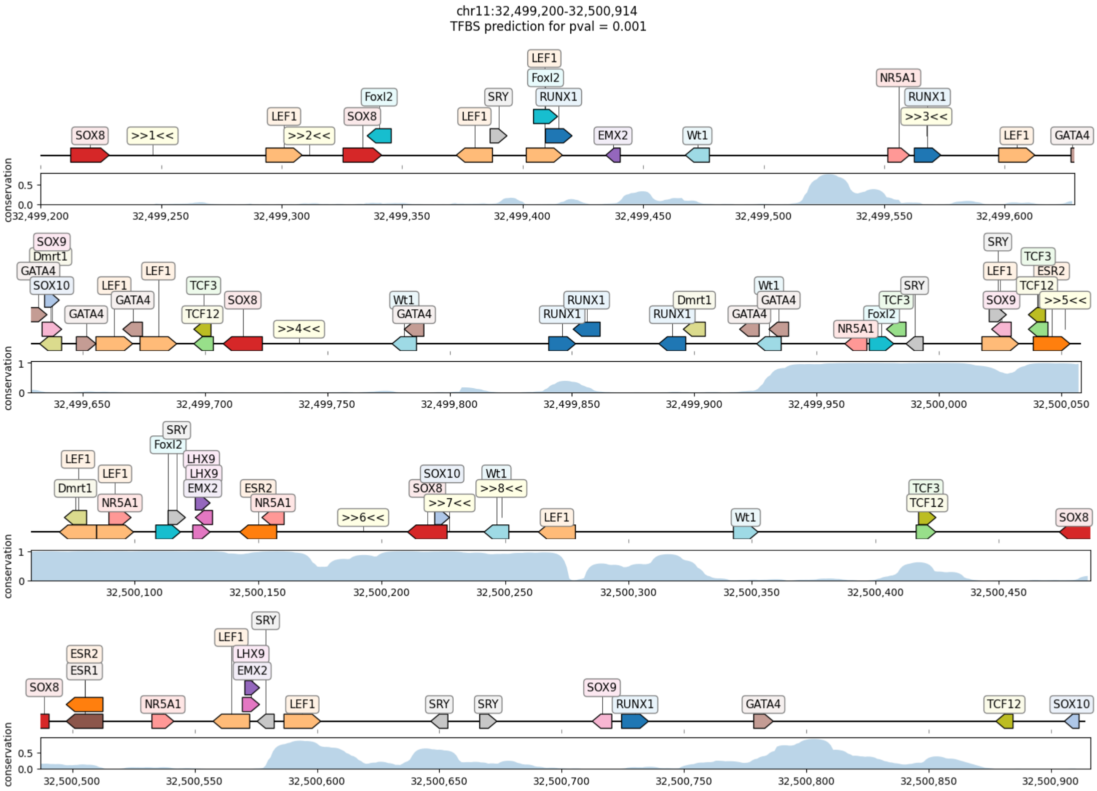

**Supplementary Figure 8. Rare variants within the human conserved *WT1* upstream enhancer identified in a DSD cohort.** (A) Summary of rare non-coding variants located within the homologous *WT1* upstream enhancer found accessible in human ATAC-seq data identified by either whole-genome sequencing or Sanger sequencing in a 400 patient DSD cohort. Genomic coordinates are reported according to the human reference genome (hg38). Population allele frequencies (AFpopmax) were obtained from gnomAD. Distance indicates the number of base pairs between the variant and the *WT1* transcription start site. Inheritance status is shown when parental data were available. Phenotype and reported ancestry are indicated for each individual. Variants classified as novel were absent from public population databases at the time of analysis. Abbreviations: Chr, chromosome; Ref, reference allele; Alt, alternate allele; AFpopmax, maximum allele frequency (gnomAD); TAD, topologically associating domain; bp, base pairs; –, not available. Distances are given in bp. (B) Predicted transcription factor binding sites within the human *WT1* enhancer region harboring candidate variants identified in individuals with DSD. The genomic interval is shown with predicted transcription factor binding sites identified using JASPAR motif analysis ($p \leq 0.001$). Candidate variants are indicated along the sequence together with predicted binding motifs for transcription factors involved in gonadal development and sex determination. The conservation track below each segment indicates evolutionary conservation across species. Coordinates are shown relative to the human reference genome (hg38).

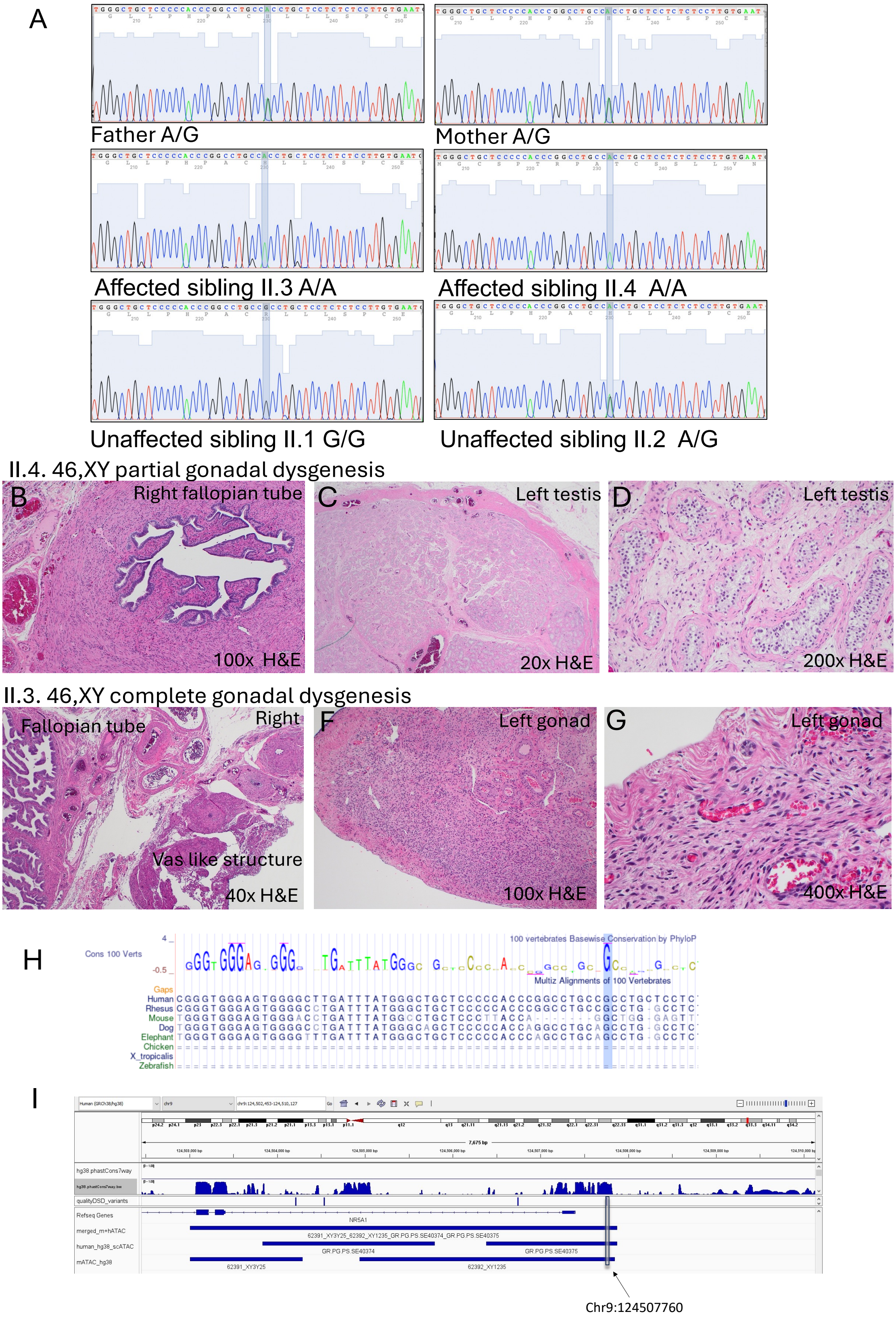

**Supplementary Figure 9. Variant located upstream of the *NR5A1* gene segregates with DSD in a pedigree.** (A) Sanger sequencing around the G>A variant located 361 bp upstream to the *NR5A1* gene. Parents and unaffected sibling are heterozygous, another unaffected sibling is G/G and two affected 46,XY DSD siblings are A/A. (B) Histology images from affected individual II.4 showing a fallopian tube on the right side (which also had a streak gonad – not shown), and a dysgenic testes on the left (C,D). (E) Histology from affected individual II.3 showing the presence of both fallopian tube and vas deferens-like structures on the right, and a dysgenic gonad with ovarian like stroma on the left (F,G). (H) Conservation of the variant among various mammals and non-mammalian species. (I) The variant, located 361 bp upstream to the human *NR5A1* gene, is located in a highly conserved and accessible region in human and mouse.

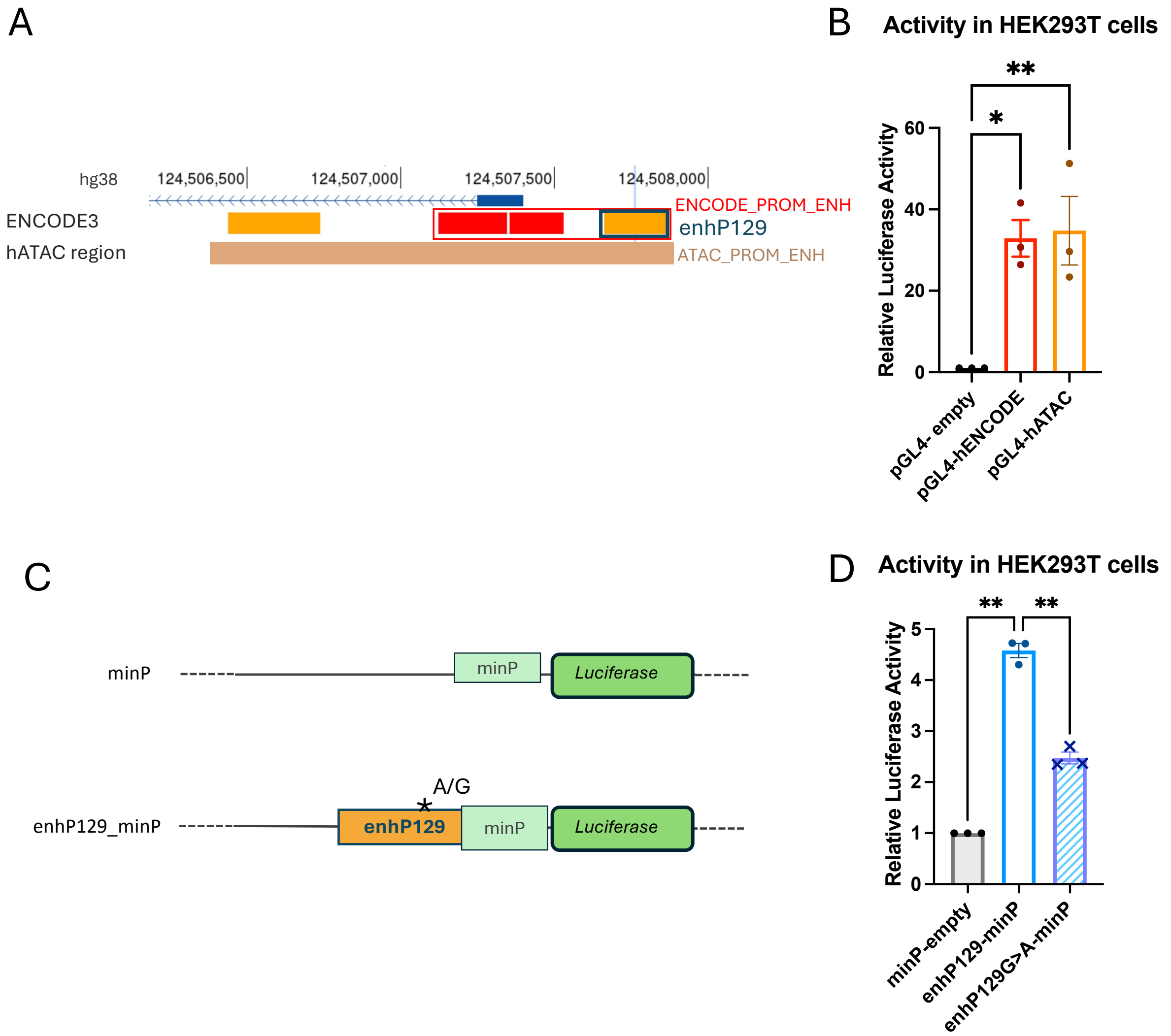

**Supplementary Figure 10. Luciferase assays using the putative enhancer upstream of *NR5A1*.**

(A) Schematic showing the cloned enhancer/promoter regions. (B) The relative luciferase activity of each *NR5A1* enhancer region (normalised against a Renilla luciferase transfection control and relative to the empty vector) in HEK293T cells. (C) A schematic indicating the proximal enhancer (enhP129 corresponding to EH38E2726129), containing the G or A variant, cloned into a luciferase plasmid upstream of a minimal promoter. (D) Luciferase activity of this enhancer fragment with and without the patient variant, relative to the empty minP control vector in HEK293T cells. The variant causes a 46% reduction in activity on average. Each data point represents the mean from three technical replicates, with three biological replicates (independent experiments) shown. Bars are SEM. P-values calculated with ANOVA *** <0.001, ** <0.01, * <0.05.

| Table S1. Primers for genotyping mice | | | | |
|---|---|---|---|---|
| **Target** | **Primer name** | **Description** | **Sequence 5' to 3'** | **Product and size** |
| Sex | Sex_F | X/Y chromosome | GATGATTTGAGTGGAAATGTGAGGTA | Sex_F + Sex_R: 280bp in XY; 685bp,660bp and 480bp in XX (McFarlane et al., 2013) |
| | Sex_R | X/Y chromosome | CTTATGTTTATAGGCATGCACCATGTA | |
| *Wt1_Enh* External | *Wt1* _ups_F1 | Flanks 5' end of *Wt1_Enh* | GTATAACCCTGAGTCTGGAC | *Wt1* _ups_F1+*Wt1* _ups_R1: 1330 bp in wild type allele, 410 bp in del allele (931 or 934 bp deletion) |
| | *Wt1* _ups_R1 | Flanks 3' end of *Wt1_Enh* | GGTTATAGGTACATCTCACCTAG | |
| *Wt1_Enh* Internal | *Wt1* _ups_Internal F1 | Internal to the *Wt1_Enh* region | CTGAGAAGCCTGAAGAATG | Wt1_ups_Internal F1+*Wt1* _ups_R1: 732 bp in wild type allele, no band in deleted allele (931 or 934 bp deletion) |
| | *Wt1* _ups_R1 | Flanks 3' end of *Wt1_Enh* | GGTTATAGGTACATCTCACCTAG | |

| Table S2. sgRNA used for CRISPR genome editing | | |
|---|---|---|
| **sgRNA ID** | **Target Sequence** | **PAM sequence** |
| *Wt1* _ups_G1_5' | GAGCCTAATAGAGTGGGTGA | AGG |
| *Wt1* _ups_G1_3' | CATCTTAAGAAGGATCCACA | AGG |

| Table S3. Antibodies/ Dyes used in this study | | | | | | | |
|---|---|---|---|---|---|---|---|
| **Target** | **Host** | **Manufacture** | **Cat. number** | **Dilution** | **Antibody type** | **Used for staining** | **Cell type** |
| αSOX9 | Goat | R&D systems | AF3075 | 1:300 | Primary | Adult gonads (6W), Embryonic gonads (E13.5) | Sertoli cells |
| αFOXL2 | Rabbit | Abcam | ab246511 | 1:300 | Primary | Adult gonads (6W), Embryonic gonads (E13.5) | Granulosa cells |
| αGCNA1 (TRA98) | Rat | Abcam | ab82527 | 1:200 | Primary | Adult gonads (6W), Embryonic gonads (E13.5) | Germ cells |
| αSF1 | Rat | Cosmo Bio | KAL-KO610 | 1:300 | Primary | Adult gonads (6W), Embryonic gonads (E13.5) | Sertoli and Leydig cells |
| αWT1 | Rabbit | Santa Cruz | sc-192 | 1:100 | Primary | Adult gonads (6W), Embryonic gonads (E13.5) | Sertoli, granulosa, IM |
| αGATA4 | Mouse | Santa Cruz | sc-25310 | 1:300 | Primary | Adult gonads (6W), Embryonic gonads (E13.5) | Early Sertoli cells |
| αMouse-Alexa flour 647 | Donkey | Invitrogen | ab150111 | 1:500 | Secondary | Adult gonads (6W), Embryonic gonads (E13.5) | - |
| αGoat-Alexa flour 488 | Donkey | Invitrogen | A-11055 | 1:500 | Secondary | Adult gonads (6W), Embryonic gonads (E13.5) | - |
| αRabbit-Alexa flour 568 | Donkey | Invitrogen | A-10042 | 1:500 | Secondary | Adult gonads (6W), Embryonic gonads (E13.5) | - |
| αRat-Alexa flour 647 | Donkey | Abcam | ab150155 | 1:500 | Secondary | Adult gonads (6W), Embryonic gonads (E13.5) | - |
| αRat-Alexa flour 488 | Donkey | Abcam | AB150153 | 1:500 | Secondary | Adult gonads (6W), Embryonic gonads (E13.5) | - |
| αMouse-Alexa flour 568 | Donkey | Invitrogen | A10037 | 1:500 | Secondary | Adult gonads (6W) | - |
| αRabbit-Alexa flour 647 | Donkey | Abcam | ab150067 | 1:500 | Secondary | Adult gonads (6W) | - |
| DAPI | - | Invitrogen | D-1306 | 300nM | Secondary | All | Nucleic acid |

| Table S4. Primers used for quantitative RT-PCR | | | |
|---|---|---|---|
| **Primer name** | **Gene** | **Marker of** | **Sequence 5' to 3'** |
| *Sox9* F | *Sox9* | Sertoli cells | AAGAAAGACCACCCCGATTACA |
| *Sox9* R | | | CAGCGCCTTGAAGATAGCATT |
| *Sox8* F | *Sox8* | Sertoli cells | AGCGAGAAGAGGCCGTTTG |
| *Sox8* R | | | TCAGTACCAGAGTCTGAGTCG |
| *Hsd3b* F | *Hsd3b* | Leydig/techa cells | CTCAGTTCTTAGGCTTCAGCAATTAC |
| *Hsd3b R* | | | CCAAAGGCAAGATATGATTTAGGA |
| *Cyp11a1* F | *Cyp11a1* | Leydig/techa cells | CACTTCTGGAGGGAGAGTGG |
| *Cyp11a1* R | | | ATGCCTGGAAGAAAGACCGA |
| *Nr5a1* F | *Nr5a1* | Steroidogenic marker | CCTCGATGTGAAATTCCTGAACA |
| *Nr5a1* R | | | TCCTGGGCGTCCTTTACG |
| *Foxl2* F | *Foxl2* | Granulosa cells | CGGCATCTACCAGTACATCATAGC |
| *Foxl2* R | | | GCACTCGTTGAGGCTGAGGTTG |
| *Rspo1* F | *Rspo1* | Granulosa cells | GGGATCAAGGGCAAGAGACAG |
| *Rspo1* R | | | CTGGCGGATGTCGTTCCTC |
| *Nr2f2* F | *Nr2f2* | Stromal cells (Ovary) | TCAACTGCCACTCGTACCTG |
| *Nr2f2* R | | | CCATGATGTTGTTAGGCTGCAT |
| *Wt1* F | *Wt1* | Sertoli/Granulosa cells | TTGAATGCATGACCTGGAATCA |
| *Wt1* R | | | TTCCCTTTAAGGTAGCTCCTAGGTT |
| *Gata4* F | *Gata4* | Sertoli/Granulosa cells | CCCCAATCTCGATATGTTTGATG |
| *Gata4* R | | | TTGACACACTCTCTGCCTTCTGA |
| *Hprt* F | *Hprt* | House keeping gene | GCTTGCTGGTGAAAAGGACCTCTCGAAG |
| *Hprt* R | | | CCCTGAAGTACTCATTATAGTCAAGGGCAT |

| Table S5. Primers used for Sanger sequencing and Luciferase Assay cloning | | | | | |
|---|---|---|---|---|---|
| **Name** | **primer sequence 5-3'** | **Primer detail** | **Product size** | **Vector backbone/hATAC peak** | **hg38. annotation** |
| NR5A1_9-124507760-G-A_for | CCTTCTTTGTTGGTGTTTCT | Sanger sequence | | n/a | hg38. chr9:124507499-124507848 |
| NR5A1_9-124507760-G-A_rev | CCTGACTCTACTCCAATGTC | Sanger sequence | 350bp | n/a | hg38. chr9:124507499-124507848 |
| 1F_hWT1_intron_enh | TCATGATCAGAACAAGAACTAGGC | Sanger sequence | | 64976_XY1235_GR.PG.PS.SE6706 | chr11:32430326-32431840 |
| 1R_hWT1_intron_enh | GGGAAGCTTCTCTTCCACCT | Sanger sequence | 781bp | 64976_XY1235_GR.PG.PS.SE6707 | chr11:32430326-32431841 |
| 2F_hWT1_intron_enh | GGAACATATGAAAGGCAGAGC | Sanger sequence | | 64976_XY1235_GR.PG.PS.SE6708 | chr11:32430326-32431842 |
| 2R_hWT1_intron_enh | GCTGGCGGATTTATGTCAGT | Sanger sequence | 784bp | 64976_XY1235_GR.PG.PS.SE6709 | chr11:32430326-32431843 |
| 3F_hWT1_intron_enh | AAGGACGCTGGTACCTTCAA | Sanger sequence | | 64976_XY1235_GR.PG.PS.SE6710 | chr11:32430326-32431844 |
| 3R_hWT1_intron_enh | AGACCAAGGCTGTGTGTGTG | Sanger sequence | 829bp | | |
| NR5A1_ENC127-129_F | GGTGGTGATATCGAGGGTGAGTCTGGGAGA | Luciferase promoter assay | 844bp | pGL4.20 luc2 (cloning sites, EcoRV,/XhoI), no minimal promoter | hg38. chr9:124507097-124507940 |
| NR5A1_ENC127-129_R | GGTGGTCTCGAGCCTCTTCTTCCACCTGATG | Luciferase promoter assay | 844bp/1722bp | pGL4.20 luc2 (cloning sites, EcoRV,/XhoI), no minimal promoter | hg38. chr9:124507097-124507940/chr9:124506219-124507940 |
| NR5A1_hATAC40375_F | GGTGGTGATATCTCACTTCTCCTCTCTGAAAC | Luciferase promoter assay | 1722bp | pGL4.20 luc2 (cloning sites, EcoRV,/XhoI), no minimal promoter | hg38. chr9:124506219-124507940 |
| NR5A1_EnhP129_F | GTTGTTGATATCGAGCCCAGAGGCAGGGA | Luciferase enhancer fragment assay | | pGL4.26 (luc2,minP) (cloning sites, EcoRV,/XhoI), minP minimal promoter | |
| NR5A1_EnhP129_R | GTTGTTCTCGAGCTTGCCTCCGCACTGG | Luciferase enhancer fragment assay | 216bp | pGL4.26 (luc2,minP) (cloning sites, EcoRV,/XhoI), minP minimal promoter | |

| Table S6. Vectors used for Luciferase Assays | | |
|---|---|---|
| Vector name | Company | purpose |
| pGL4.10[luc2] | Promega (E6651) | a basic reporter vector encoding the luciferase gene luc2 (Photinus pyralis) and does not contain no promoter. (to analyse promoter elements) |
| pGL4.26[luc2/minP/Hygro] | Promega (E8441) | a reporter vector encoding the luciferase gene luc2 (Photinus pyralis) and a minimal promoter (to analyse response elemtents) |
| pRL-TK (Renilla) | Promega (E2241) | Control reporter vector, constitutive expression of Renilla under the control of the HSV-thymidine kinase promoter. |
